# The association between cholera and weather: a systematic review and direction-of-effect meta-analysis

**DOI:** 10.1101/2025.11.04.25339479

**Authors:** Qulu Zheng, Andrew S. Azman, Elizabeth C. Lee, Rebecca Shade, Gina E. C. Charnley, Flavio Finger, Raoul Kamadjeu, Benjamin Zaitchick, Javier Perez-Saez

## Abstract

**Background:** Cholera is often used as a model of the effect of climate and weather on infectious diseases. Yet the empirical evidence remains inconsistent, and the few existing reviews have not provided a synthesis of evidence nor systematically assessed sources of heterogeneity. Our objective was to address both of these gaps.

**Methods:** We conducted a systematic review on the association between cholera and precipitation, temperature, floods and drought. PubMed, Scopus, Global Health, and ProQuest were searched to identify studies before 18 May, 2023. We extracted data on exposures, outcomes, and analytical methods, along with contextual study factors, and assessed study reporting completeness. Evidence synthesis focused on the direction of association using the vote counting method, and we performed a direction-of-effect meta-regression analysis to attribute variability to contextual factors.

**Findings:** We included 60 studies spanning 2000-2020. Study characteristics were highly heterogeneous, with a geographical bias towards Asia (Bangladesh) vs. Africa. Only 23/60 studies had full reporting completeness scores with the most frequent limitation being lack of information on the epidemiological outcome. Among all studies, we found a positive association between cholera incidence and precipitation (p < 0.001), temperature (p < 0.001) and floods (p 0.021). When subsetting to studies with complete reporting no association remained statistically significant. The variability in association direction was primarily attributable to study-level variability, and not to contextual factors.

**Interpretation:** The current landscape of evidence on the association between cholera and weather patterns is fragmented and suffers from important geographical biases and methodological limitations. Climate effects on cholera should therefore be systematically evaluated and discussed in the context of other known drivers of cholera risk. There is an important opportunity for future studies with transparent reporting, improved data quality, and explicit consideration of contextual and epidemiological factors to generate reliable evidence to guide climate-related cholera forecasts and mitigation approaches.

**Funding:** This study was funded by the Welcome Trust.

**Research in context:** *Evidence before this study:* We conducted a PubMed search for previous reviews on August 16, 2024, with no language or date restrictions using the search terms [“cholera*” AND (“climate” OR “weather”) AND (“Systematic review” OR “narrative review” OR “science review”)]. Of the 20 reviews identified, only six examined the associations between cholera and the key weather exposures (precipitation, temperature, flood, and drought). Among these, two provided global coverage, three focused on Africa, and one on Asia. Two articles assessed all four weather exposures, while others addressed only one or two. All six reviews qualitatively summarized the association effects between weather exposures and cholera, but none synthesized the association effects systematically (e.g., the direction of the association effect). Findings across reviews were heterogeneous, with variations by setting (e.g., inland versus coastal regions). Notably, none of the reviews systematically evaluated study quality nor completeness of reporting, and only one assessed the statistical methods used to estimate associations.

*Added value of this study:* This study addresses a critical gap by providing a comprehensive synthesis of the evidence linking key weather exposures to cholera outcomes, including a quantitative assessment of the direction of association effects. Additionally, this study investigates the sources of heterogeneity. We found that most of the included studies focused on Asia and Africa, with a bias towards Bangladesh (Asia), and spanned over the period from 2000 to 2020. Study characteristics, including the spatio-temporal coverage, cholera outcome definitions, and study reporting completeness, as well as the analytic methods varied widely across different studies. Consistent with previous reviews, we observed substantial heterogeneity in effect estimates. In evidence synthesis we found a statistically significant positive association among all studies between cholera incidence and precipitation, temperature and floods, but not in studies with full reporting completeness scores. Our meta-analysis further suggested that the observed variability in directions of associations was primarily driven by within-study error variance and between-study differences, rather than contextual or epidemiological factors.

*Implications of all the available evidence:* Current evidence on weather and cholera associations is highly inconsistent due to geographical biases, methodological heterogeneities and data limitations. This review highlights the need for future studies on the association between cholera and weather in cholera-affected countries that have been relatively less investigated, using improved cholera epidemiological data based on standardized definitions following established international guidelines, with transparent reporting on exposures, outcomes and analytical methods, and accounting for epidemiological and contextual factors.

## Introduction

Climate and weather have long been recognized as key factors influencing infectious disease dynamics across pathogens and transmission pathways (1–4). Understanding these associations is foundational for attributing disease burden to climate and weather drivers, and for projecting the consequences of climate change, including rising temperature, water cycle intensification, and increases in extreme weather and climate events (5–11). Cholera, a waterborne bacterial pathogen, is frequently cited as a model for understanding the relationship between climate and infectious diseases in a changing climate (12).

Cholera is caused by the ingestion of toxigenic *Vibrio cholerae* (typically serogroup O1), which remains a global health threat with more than 535,000 suspected cases and 4,000 reported deaths in 2023 (13,14). Evidence on the link between cholera and climate dates back to the 19th century where outbreaks in the Indian subcontinent were associated with the rainy season (15,16). These early observations were supported by seminal studies on the association between cholera and the El Nino Southern Oscillation (ENSO), a climate anomaly known to modulate seasonal monsoon patterns (17–19). The link between climate and cholera in turn implies a mechanistic effect of local weather patterns that characterize climate and its anomalies on disease transmission. Multiple mechanistic hypotheses for how cholera transmission occurs have been proposed, which each imply different potential pathways for climate and weather effects. One hypothesis originated in the Indian subcontinent, dubbed the “cholera paradigm”, suggests that cholera burden is in part driven by exposures to naturally occurring *V. cholerae* reservoirs in environmental water and that the growth and persistence of these bacteria is modulated by weather through its effects on aquatic habitats (20). A second hypothesis is that cholera is mainly driven by proximal human-to-human routes involving the household and/or local environments, with spatial dispersal occurring through human mobility, this hypothesis having the strongest support from genomic evidence (21,22). In this latter hypothesis, seasonal weather patterns and anomalies may impact cholera transmission by compromising safe water, sanitation and hygiene (WASH) access, altering human mobility and/or exacerbating underlying risk factors (23,24).

The link between cholera and climate has received increasing interest in the past 20 years (12,25,26). For instance, the World Health Organization (WHO) mentioned climate change as a cause of high cholera incidence rates in 2023-2024 that led to the declaration of a global cholera emergency (27). Cholera also features prominently in policy reports on climate change adaptation by the Intergovernmental Panel on Climate Change (28), and cholera’s climate sensitivity is referenced by the Global Task Force on Cholera Control (GTFCC) for strategic recommendations on targeting public health interventions, such as WASH improvements or the preventive use of limited supplies of oral cholera vaccine in cholera-affected countries (29). However, despite this growing policy interest, a synthesis of evidence on the direction and strength of cholera’s associations with weather patterns and anomalies remains lacking, which severely limits our understanding of the overarching link between cholera and climate across settings.

Previous literature reviews on the associations between cholera and weather patterns have found largely variable results (30–37), that can be attributed to two main sources of heterogeneity. The first is the complex and dynamic nature of cholera transmission, yielding different mechanistic relationships with weather across climatic and epidemiologic settings (36). For instance, the association between rainfall and cholera in the Indian subcontinent has been found to be negative before the monsoon season but positive after it, with various mechanistic explanations ranging from seasonal changes in environmental water chemistry to WASH disruption (35,36). Similarly, there are apparently conflicting hypotheses related to floods. Floods drive population displacement and WASH infrastructure disruption, so they are hypothesized to increase cholera incidence, but dilution and washout of bacteria in surface waters during floods is hypothesized to decrease cholera incidence (38). In contrast, cholera burden has been consistently posited to be exacerbated by high temperatures and droughts by favouring bacterial concentrations through persistence and growth, and by increasing risk due to malnutrition and limitations in clean water supply and safe cooking conditions (23,33).

The second source of heterogeneity is variability in study methodology, including limited high-quality cholera and climate/weather data at matching spatio-temporal scales, especially in resource-limited regions, variability in epidemiological data quality, including inconsistent cholera case definitions and inherent differences in surveillance system performance, differences in epidemiological outcomes, and differences in study designs and statistical methods (30,33,35,37). It is thus difficult to assess the reliability and comparability of findings across studies, and whether differences in published measures of association between cholera and weather variables arise from true setting-specific epidemiological differences or from analytical artifacts. As a result, a synthesis of evidence is yet unavailable regarding the strength or even the direction of associations between cholera and often-cited weather variables and anomalies, chief among which are precipitation, temperature, floods and droughts.

In this study we synthesize existing evidence on cholera and weather through a systematic review and meta-analysis of the direction of association between cholera and key weather variables and their anomalies. We focus on precipitation, temperature, floods and droughts, and systematically evaluate known sources of study heterogeneity, including study settings, exposure and outcome data, and analytical approaches. We then synthesize evidence on the direction of associations, and meta-analyze their variability across possible sources of heterogeneity.

## Methods

### Systematic review

#### Search strategy

We conducted a comprehensive search of four databases: PubMed, Scopus, Global Health, and ProQuest Science on September 22, 2021 for titles and abstracts with the following search terms: “(cholera*) AND ((climate) OR (weather) OR (rain*) OR (temperature) OR (precipitation) OR (flood*) OR (drought) OR (humidity) OR (moisture))”. We further updated this search on May 18, 2023 using the same search strategy.

#### Screening and full-text review

After removing duplicate entries, two independent reviewers screened all titles and abstracts for inclusion based on specific criteria. Inclusion criteria at the screening step included epidemiologic studies of weather, climate, and cholera in humans. Exclusion criteria at the screening step included modeling studies lacking primary data, reviews, abstracts, commentaries, laboratory and animal studies, and studies that did not focus on pandemic serogroups *V. cholerae* El Tor O1 and O139. Additionally, studies that examined the occurrence of *V. cholerae* in aquatic environments (e.g., natural waters) without epidemiologic considerations, or those focusing solely on epidemiologic aspects without weather or climate considerations were excluded. During the full-text review, additional exclusion criteria included a lack of original data, duplicated data, and irrelevant exposures. Studies were also excluded if they asserted a relationship between relevant exposures and outcomes without providing measures of association, did not focus on cholera as an outcome, or were inaccessible. After full text review, we focused data extraction on studies looking into at least one of four main weather exposures of interest: precipitation, temperature, floods and droughts.

#### Data extraction

For each study, we extracted the information relevant to the association effects of selected climatic variables i.e., precipitation, temperature, flood, and drought, on cholera. Some studies reported multiple association effects, all of which were extracted. We gathered a comprehensive set of variables at both the study and on analytical methods. Study-level variables include the timeline of the study (lower and upper bounds), the country where the study was conducted, and any mechanistic explanations provided by authors. The analytical methods variables include exposure measurement, outcome measurement, association effect measurement, and others. For each exposure measurement, we extracted the exposure category, (precipitation, temperature, flood or drought) temporal and spatial scales, and data source. For each outcome measurement, we extracted the outcome category (main ones were incidence and occurrence), case definitions, whether the cholera cases were medically attended, and temporal and spatial scales. The variables extracted for each association measurement included the category of the association effect, effect size, the function used to transform the association effect if any, whether the analysis controlled for confounding non-exposure variables (e.g., WASH), the direction of effect, measures of uncertainty, and the statistical method used to assess this association measurement.

Additionally, we noted whether this association measurement was part of primary or sensitivity analyses. Association effect categories include ratio, correlation coefficient, risk difference, attributable risk, and others. Other variables extracted included the geographic information of the association effect, the reporting period for outcome cases (lower and upper bounds), the lag time between exposure and outcome measurements, and whether the spatial resolutions of exposure and outcome measurements were aligned.

#### External data extraction

To evaluate whether the heterogeneity in association effect is attributable to study-specific factors or epidemiological determinants, we incorporated key epidemiological variables thought to influence cholera transmission into our analysis. These variables include a location’s proximity to the ocean (inland vs. coastal), urbanicity, frequency of cholera reporting to the WHO, climatic zone, and the average Human Development Index (HDI) as a proxy for WASH conditions. Details on the methods for the definition of these variables are given in Supplementary Materials (page 2).

### Data analysis

#### Assessing study reporting completeness

Due to the variability in study designs and methodological approaches, we did not attempt to evaluate overall study quality following established guidelines but instead focused on assessing the degree to which studies reported minimal elements to understand study exposures and outcomes and to allow for study reproducibility. To evaluate the completeness of reporting for each study, we developed a score system based on nine variables covering three main dimensions (Supplementary Table S14): the exposure (climate/weather variable), the epidemiologic outcome, and the measure of association. Each variable was assigned a score of 1 if the study provided specific and clear information relevant to that variable, and a score of 0 if it did not. For each study, we calculated a total reporting score and three component scores, including the exposure-, outcome-, and association-related reporting completeness score for every exposure and outcome association pair by summing the scores across all the corresponding variables. When multiple estimates were reported for the same exposure and outcome pair (e.g. precipitation and cholera incidence but at different spatial scales), we assigned that pair the minimum score across estimates. Studies were considered to meet minimal reporting criteria when they had a full score of 9out of 9 questions.

#### Synthesis of evidence

Given the strong heterogeneity in how studies defined exposures, outcomes, and in statistical methods, we chose to synthesise results in terms of the direction of associations between cholera and weather exposures using the vote counting method (39), stratifying by type of cholera outcome. As each study may report multiple estimates for each exposure/outcome pair, the direction of association between each weather exposure and cholera outcome was assessed using the percentage of effects in either direction following established methods for direction of effect plots, resulting in studies being classified as either “positive”, “negative”, or “conflicting” (absence of consensus direction of association in study estimates) (40). To assess the statistical significance of directions of association we used the sign test with a two-sided p-value at a 5% significance level against the null, excluding studies that had conflicting results following previous studies (41). In addition, we computed the p-values from a trinomial test to incorporate studies with conflicting results at a 5% significance level (42).

#### Meta-regression of directions of effect

We did not perform effect size meta-analysis due to the large heterogeneity in study characteristics. We instead performed a meta-analysis of direction of effect for each weather exposure-cholera outcome combination. We did so using a hierarchical Bayesian logistic regression model with a study level random effect, incorporating study characteristics including study location, factors that have been posited to be mechanistically mediated the relation between weather, climate and cholera, and analytical methods. Contextual factors included continent, climate zone, urbanicity, rainy season, inland vs. coastland, Human Development Index, and frequency of cholera reporting to WHO. Analytical factors included lag between exposure and outcome and the temporal scale of outcome (see Section “External data extraction”). We focused this analysis on studies at national or subnational scales for which contextual factors could be defined. A full description of the direction-of-effect meta-analysis is given in Supplementary Material section 2.

We performed inference using a Bayesian hierarchical model with a Hamiltonian Monte Carlo sampler as implemented in the Stan programming language (43). To account for missing covariate values, we marginalized over possible missing covariate levels based on observed frequencies of study characteristics (44).

### Software and code

We utilized Covidence software for the screening and full-text review process (https://www.covidence.org/). All extracted data and data processing code are publicly available on Github at https://github.com/HopkinsIDD/cholera_climate_review.

## Results

Our literature search yielded 3,794 results, of which 2,927 were screened after removal of duplicates (Figure 1). A total of 361 studies were assessed for eligibility. We excluded 301 studies that did not meet eligibility criteria, the most frequent (mutually-exclusive) reasons for exclusion being not presenting original data (N=87), not reporting a statistical measure of association (N=86), or reporting on an irrelevant exposure (N=85).

**Figure 1.**
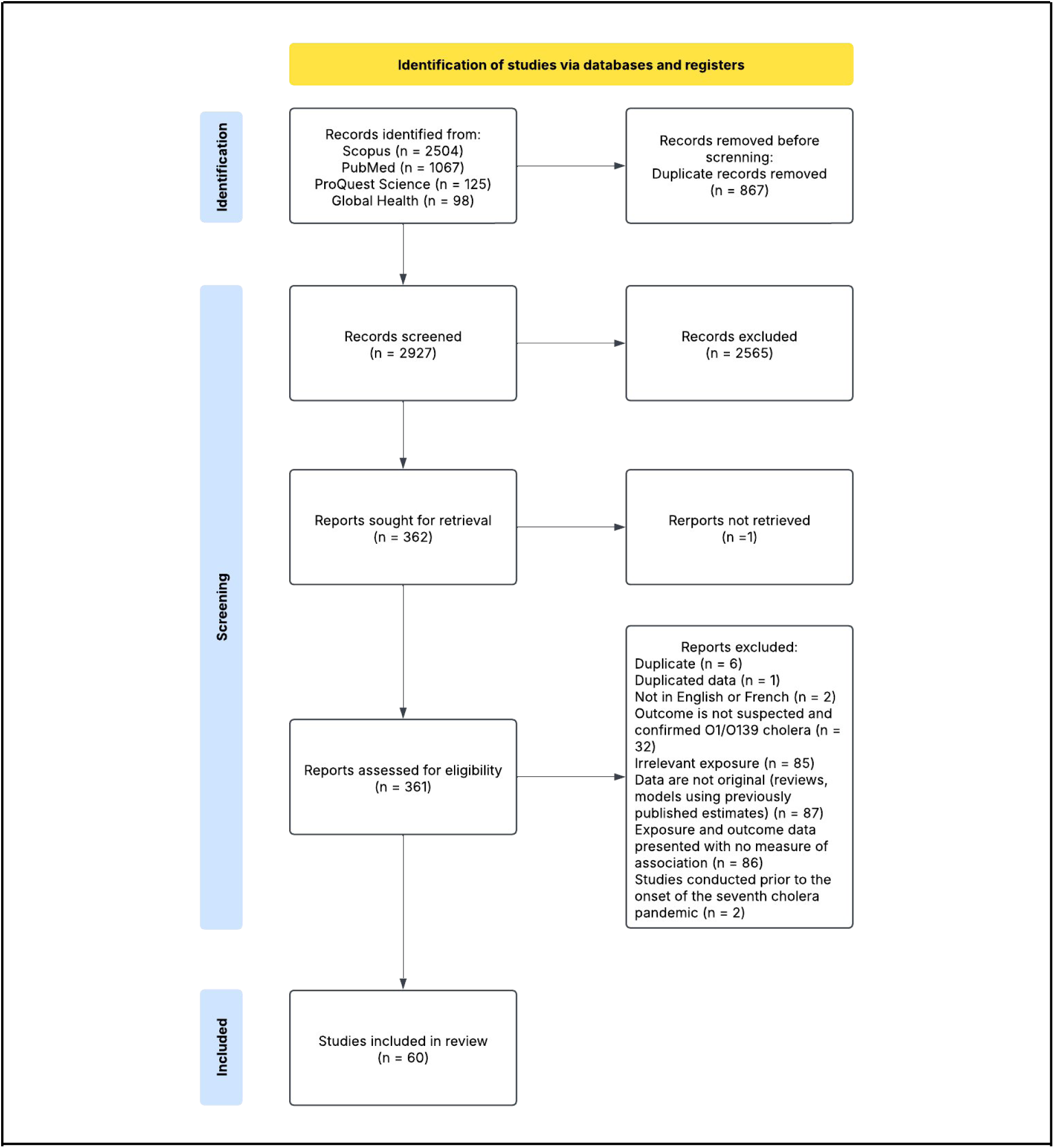
PRISMA flow diagram.

Our analysis dataset included 60 studies that met eligibility criteria. Most of these studies focused on the continents of Asia (N=27) or Africa (N=20) at subregional (national or subnational) levels (Figure 2a, Supplementary Figure S1), and the majority covered the period 2000-2020 (Figure 2b), with similar study durations (Supplementary Figure S2).

**Figure 2.**
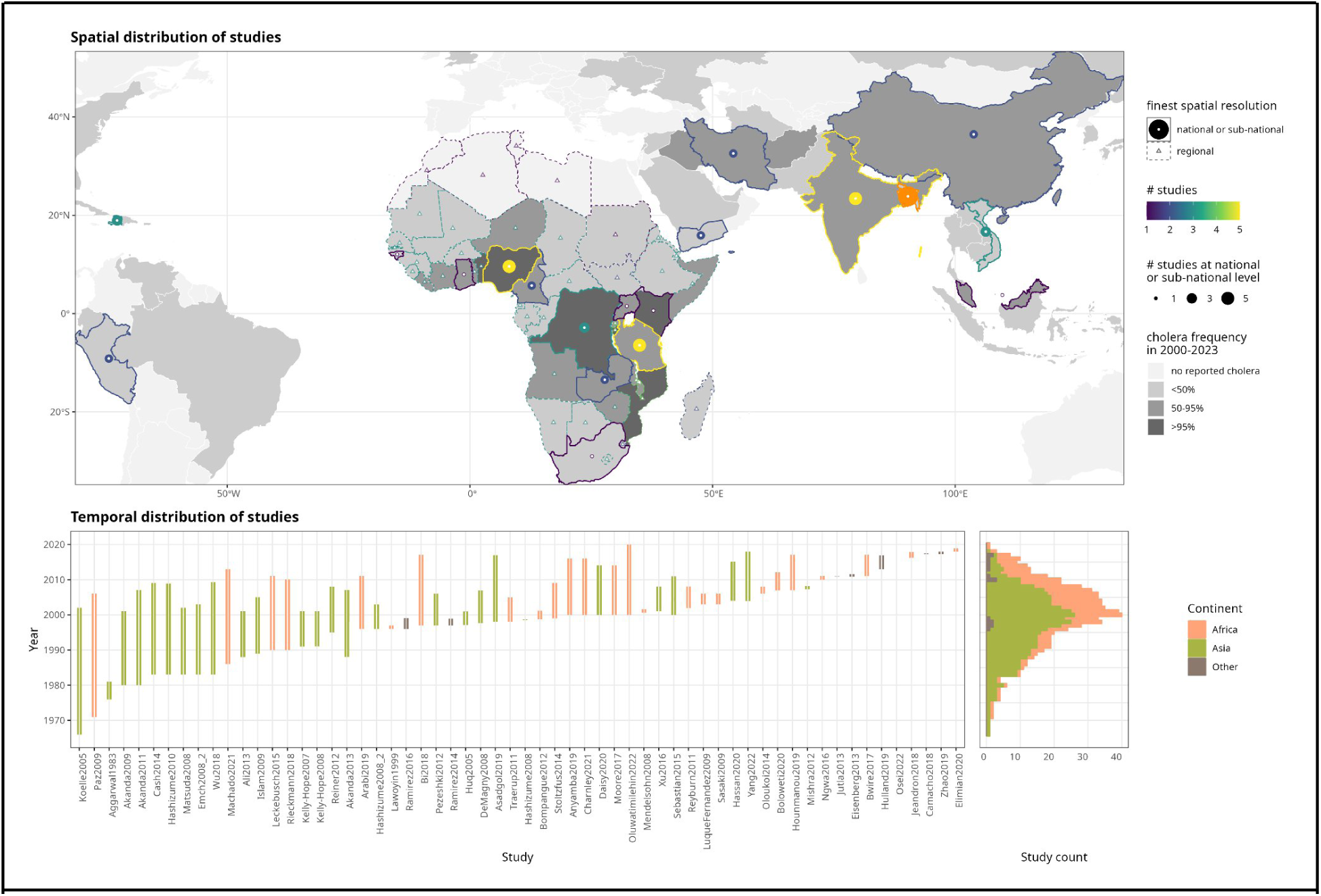
Spatio-temporal coverage of studies on cholera and weather. Top) Spatial distribution of retained studies. Colored country borders indicate countries covered by at least one regional (dashed) or national/sub-national (full line) study, with border color indicating the total number of covering studies. National and subnational-level studies are indicated with dots, with size and color indicating the number of studies (orange = 16 studies in Bangladesh). Country fill (grays) indicates the fraction of years in 2000-2023 that the country reported at least one cholera case to the WHO. Bottom) Temporal coverage. Left) Bars give the start and end years of each study, shown by continent (fill color). Right) Number of studies covering each year by continent (fill color).

Among the 48 countries in Africa with reported cholera to WHO between 2000 and 2023, only 21% (N=10) were covered by at least one study at national or subnational level, as opposed to 100% (N=6 of 6) in Asia. Notably, Bangladesh (Asia) was the single country with most studies at subnational levels (N=16), although studies were primarily concentrated in two specific locations (Dhaka and Matlab), and for which among the longest time periods were covered (Supplementary Figure S3).

We found large variability across multiple study dimensions, including spatio-temporal coverage, outcome definition, statistical methods to quantify associations, and study reporting completeness (Table 1, Supplementary Table S1). Most studies reported on the association between cholera and precipitation (N=51) or temperature (N=30), with fewer studies addressing floods (N=11) or droughts (N=4). The main type of reported outcome was cholera incidence (i.e., the number or rate of reported cholera cases, N=49), with few studies reporting on occurrence (i.e., the presence/absence of reported cholera cases, N=5), or other types of outcomes (Supplementary Table S2). Only 36/60 (60%) studies provided information on the study population, most of which were medically-attended cases (N=30). Nearly half of the studies did not mention a cholera case definition (N=24). Among those that did, few used microbiologically-confirmed cases only (N=15), the rest using at least in part suspected cholera cases, commonly defined by acute watery diarrhoea (45), or other types of individual-level outcomes (Supplementary Table S2). Around half of studies used monthly epidemiologic outcomes (N=28), with fewer considering weekly outcomes (N=14). The majority of studies had aligned outcome and exposure temporal resolutions (88%, N=53), but less than half of the studies (N=27) had matching spatial scales, of which 25 had both matching spatial and temporal scales. When stratifying by continent we found similar patterns in coverage of exposure variables and in types of outcome, as well as in heterogeneity of cholera case definitions, with studies in Africa more frequently lacking a case definition than in Asia (48% vs. 31%) (Supplementary Table S3-S5). The methods used to characterise the association between cholera and weather exposures varied widely between studies. The two most commonly used statistics were correlation coefficients (N=32) and risk/odds ratios (N=30), with fewer studies using other metrics (Supplementary Tables S1-2).

**Table 1.** Characteristics of studies included in the systematic review (N=60). Full descriptive table given in Supplementary Table S1. Details on characteristics reported as “Other” are given in Supplementary Table S2. Note that the sum in each category may exceed the number of studies because a single study can report on multiple variables (e.g. multiple weather exposures).

| Category | Characteristic | N (%) |
| --- | --- | --- |
| **Weather exposure** | Precipitation | 51 (85%) |
|  | Temperature | 30 (50%) |
|  | Flood | 11 (18%) |
|  | Drought | 4 (67%) |
| **Epidemiologic Outcome type** | Incidence | 49 (80%) |
|  | Occurrence | 5 (8%) |
|  | Other | 7 (11%) |
| **Temporal resolution of outcome** | 1-14 days | 14 (23%) |
|  | 15-30 days | 28 (46%) |
|  | 31 days-1 year | 13 (21%) |
|  | > 1 year | 3 (5%) |
|  | Missing | 3 (5%) |
| **Cholera case definition** | Suspected | 3 (5%) |
|  | Confirmed | 15 (23%) |
|  | Deaths | 1 (2%) |
|  | Mixed<sup>1</sup> | 21 (33%) |
|  | Missing | 24 (38%) |
| **Statistical measure of association** | Risk Ratio | 30 (42%) |
|  | Correlation coefficient | 32 (45%) |
|  | Risk difference | 6 (9%) |
|  | Attributable risk | 2 (3%) |
|  | Other | 1 (1%) |

<sup>1</sup> Unspecified mix of suspected and confirmed cases

When assessing reporting completeness we found that less than a third of studies met all minimal reporting criteria (N=23, Table 2). Among studies with imperfect reporting, most lacked description of the cholera case definition (N=24), and a smaller proportion lacked critical information on the exposure (N=13) or on the statistical methods to quantify associations (N=11). Study-level scores are given in Supplementary Figure S4.

**Table 2.** Scoring of study reporting completeness. The number of studies meeting each criteria are presented by question category (exposure, outcome and association). Study-level scores are given in supplementary Figure S4. The detailed explanations on each question are given in supplementary Table S14.

| Question Category | Sub-category | N that met criteria (%) |
| --- | --- | --- |
| Exposure | Q1. Data source reported | 55 (92%) |
|  | Q2. Spatial scale reported | 52 (87%) |
|  | Q3. Temporal scale reported | 54 (90%) |
|  | **Overall exposure-related reporting score (max = 3)** | **47 (78%)** |
| Outcome | Q4. Case definition reported | 36 (60%) |
|  | Q5. Case presentation reported | 60 (100%) |
|  | Q6. Spatial scale reported | 60 (100%) |
|  | Q7. Temporal scale reported | 60 (100%) |
|  | **Overall outcome-related reporting score (max = 3)** | **36 (60%)** |
| Association | Q8. Effect measurements reported | 60 (100%) |
|  | Q9. Uncertainty reported | 49 (82%) |
|  | **Overall association-related reporting score (max = 2)** | **49 (82%)** |
| Overall score | **Overall meeting minimal reporting criteria** | **23 (38%)** |

Given the strong heterogeneity in study characteristics and methods we did not meta-analyze effect sizes, and instead synthesized findings focusing on the direction of associations between cholera outcomes (incidence and occurrence) and weather exposures (precipitation, temperature, floods and drought) (Figure 3). Overall, studies that reported a positive exposure-outcome association had similar characteristics than those that did not (Supplementary Tables S6-S8). When considering all studies irrespective of the completeness of reporting, we found a significant positive association between cholera incidence and precipitation (p-value: <0.001), temperature (p-value: <0.001), and floods (p-value: 0.021), but not with droughts (p-value: 0.63), and none of the weather variables had a statistically significant direction of association with cholera occurrence (Supplementary Tables S9). When constraining the analysis to studies having a full reporting completeness score (N=23), none of the associations between weather exposure and cholera incidence remained statistically significant (Supplementary Tables S10). When stratifying by continent, directions of association were statistically similar between Africa and Asia for precipitation and temperature, but not for floods and droughts (Supplementary Table S11-S13).

**Figure 3.**
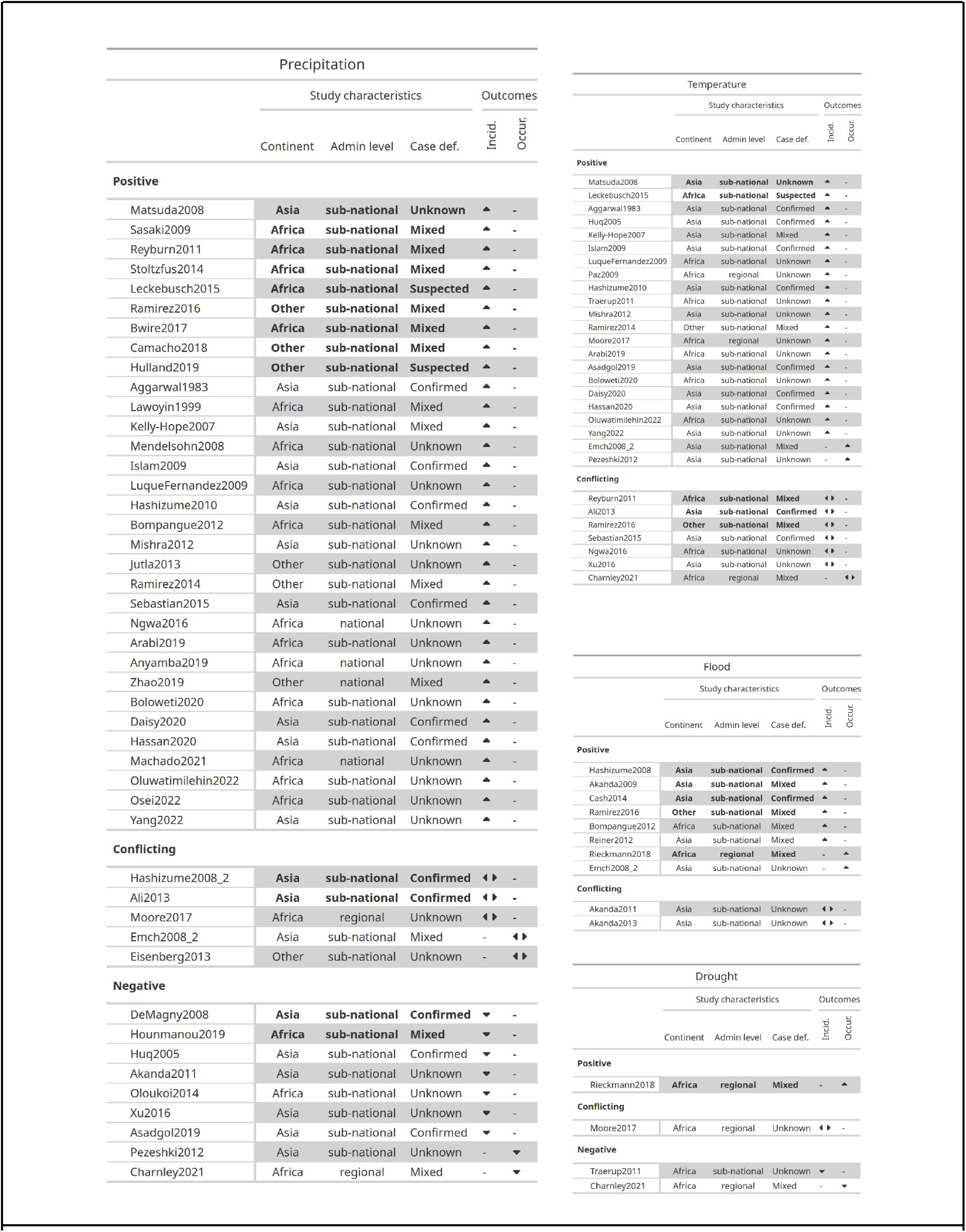
Direction of effect plot for associations between cholera and weather. The direction of association between precipitation, temperature, flood and drought are indicated by icons for the two main epidemiologic outcomes (cholera incidence and occurrence), along with selected study-level characteristics (continent, administrative level of analysis, and cholera case definition). Bold lines indicate studies with full reporting completeness score.

In direction-of-effect meta-regression analysis we found that most of the variability in association direction was due to within-study estimate error variance or between-study variability, and not to specific contextual or analytical factors (Figure S5-S6). Only one of the analytical methods we explored had a statistically significant effect, with a lower OR of positive association of cholera incidence and temperature at a lag of 3 months vs. no lag (OR 0.46, 95% CrI: 0.18-0.96). Results were similar when stratifying by continent, climatic zone and frequency of cholera case reporting (Supplementary Figures S7-S9).

## Discussion

This systematic review and meta-analysis reveals the large gap between the exemplar status of cholera as a climate-sensitive disease, and the fragmented nature of evidence linking cholera and weather. Our main finding was the strong geographical, methodological and analytical heterogeneity in existing studies linking cholera and precipitation, temperature, floods and droughts. Overall, when considering studies meeting minimal reporting requirements, evidence on the directions of association was only statistically significant for a positive effect of precipitation on cholera incidence, and inconclusive for all other weather exposures and cholera outcomes (incidence or occurrence).

Our results align with those of previous systematic reviews that have consistently highlighted the spatial and methodological heterogeneity in studies of cholera and weather/climate (23,34,37), here providing a quantitative basis for their evaluation. Among all cholera-affected countries, the majority of the evidence was generated in the Indian subcontinent, in particular in Bangladesh, especially in the cities of Dhaka and Matlab. However, Bangladesh is a unique cholera hyper-endemic area, presenting specific monsoon-driven weather patterns and ecological characteristics making Bay of Bengal a unique epidemiological setting (*46,47*). These specificities challenge the generalizability of existing evidence on cholera and weather/climate to other cholera-affected regions such as Africa and the WHO- Eastern Mediterranean regions where more than 90% of deaths were reported in 2024 (27).

On the methodological front, we find that existing evidence on the association between cholera and weather variables is based on heterogeneous definitions of exposures, epidemiologic outcomes and statistical methods used to quantify associations. Further, the majority of studies relied on data that do not meet our proposed minimum reporting standards. This is a troubling limitation for studies of cholera and weather, particularly in low resource regions, and it limits the interpretability or generalizability of findings. We note in particular the frequent lack of a cholera case definition and less than one in four studies using microbiologically-confirmed cholera cases only with the rest relying on suspected cases, echoing similar results in previous reviews (37). Suspected cholera cases typically consist of cases of acute watery diarrhea, but this definition has a low specificity, especially for younger children (48). Studies using suspected cases may therefore not accurately represent the specific sensitivity of cholera risk to meteorological conditions. For this reason, we distinguish between studies that meet minimal reporting requirements and those that do not. We however recognize that a failure to meet those requirements may stem from fundamental limitations in available data, and that our reporting completeness score is not necessarily representative of overall study quality, as studies with perfect reporting completeness scores may nevertheless present limitations in the actual data and methods used to assess associations (48).

Because of study heterogeneity we were not able to conduct an effect-size meta-analysis, and limited our synthesis of evidence to the direction of associations. We found a positive association between cholera incidence and precipitation, temperature and floods when considering all studies, but none of these associations remained statistically significant in studies with full reporting completeness scores. This synthesis only partially aligned with prior expectations based on the two main mechanistic hypotheses on cholera transmission and their implications for the role of and weather patterns, ie. exposure to *V. cholerae* bacteria in the aquatic environment, and increased risk through behavior change and WASH disruption due to severe weather events (35). For precipitation, lack of clear direction of association may have been expected as rainfall may have non-linear effects by altering transmission pathways either “concentrating” or “diluting” bacteria in the environment depending on prior hydrological conditions following the “concentration-dilution” hypothesis (36,49). The fact that we find a positive association between cholera incidence and precipitation across settings may indicate that the “concentration” effect is dominant, consistent with results for diarrheal diseases in general (49). On the other hand, the effects of temperature, floods and droughts have consistently been suggested to increase cholera incidence and occurrence due to their potential role in bacterial proliferation in aquatic environments (temperature) and on disruptions to WASH infrastructure and access (floods and droughts) (20,35). This expectation does align for temperature and floods when considering all studies, but does not for droughts, and none of the associations were significant when subsetting to studies with full reporting completeness score. Some of these discrepancies may be due to differences in study settings and effect modification due to climatic and non-climatic factors. We however did not find signals for this in our direction-of-effect meta-regression analysis as effect-level error and between-study differences dominated direction-of-effect variability.

It may be expected that cholera incidence and occurrence are differentially associated with weather variables, as the triggering of cholera outbreaks and their subsequent unfolding may be governed by mechanisms with distinct weather/climate sensitivities (50). In particular, cholera occurrence may be heavily influenced by the stochastic nature of 7th pandemic *V. cholerae* introductions into heterogeneous landscapes of population immunity in settings with differential health system response capacity, and subsequent dispersal through human mobility, as highlighted by genomic evidence of cholera dynamics across continents (51).

This dependence may dwarf the effect of weather on cholera occurrence, consistent with the lack of association found in our review. This hypothesis is however subject to testing as there were few studies to draw information from.

Elucidating the link between cholera and precipitation, temperature, floods and droughts will require future studies that meet minimal reporting requirements as the ones listed in this review, in addition to carefully addressing existing limitations on the epidemiologic quantification of cholera outcomes and their relation with weather and climate. Particular points of concern raised in this review are the need for more studies in cholera-affected regions that have received relatively less attention in the literature, particularly in Africa and the WHO-EMRO region, the use of interpretable cholera case data and the alignment of spatio-temporal scales of exposures and outcomes. We acknowledge that reliably assessing cholera incidence is an important challenge. We here stress GTFCC surveillance guidelines in the importance of confirming that cholera is circulating using traditional or rapid diagnostic tests (RDTs), and to then rigorously apply suspected case definitions (45,48). Partial case confirmation may be a valid approach when cholera outbreak outbreak occurrence is the epidemiologic outcome of interest, but may fail to adequately portray changes in incidence. For studies focusing on incidence and weather/climate associations, the gold standard would be to systematically ascertain at least a fraction of suspected cases using RDTs. With respect to the outcomes covered by the review, we found very few studies on the link between weather and cholera occurrence, as opposed to incidence, which is a gap that requires further investigation. Beyond improved macro-level studies looking into cholera and weather associations across large spatial scales, research on the link between cholera and climate will benefit from detailed micro-level case studies specific in areas with higher quality cholera and weather data, spanning geographies and epidemiologic contexts.

This systematic review comes with some limitations. First, our study quality score only accounted for reporting completeness, and we did not attempt to quantify overall study quality because of the large methodological heterogeneity and absence of established quality assessment schemes for ecological studies. Second, we were unable to provide pooled estimates of associations between cholera and weather exposures because of the same heterogeneity in analytical methods across studies. Finally, this review was limited to cover precipitation, temperature, floods and droughts, although other environmental variables have been linked to cholera in the literature such as sea surface temperature or river discharge.

We decided to focus on these variables as these were most frequently linked to cholera in previous literature reviews.

Climate change has been repeatedly cited as a cause of increases in cholera reports, which spurred the declaration of the 2023-2024 WHO global health emergency (27). This statement however stands in contrast to the results of this review that show a fragmented landscape of evidence on the link between cholera and weather with important geographical biases,methodological heterogeneities and data limitations which previous reviews had not fully systematically assessed. This highlights the importance of considering climate impacts in the context of the many other recognized drivers of cholera risk, including immunity dynamics, stochastic bacterial introductions and human-induced disruptions such as armed conflict (31,52). The results of this review also underscore the yet unexploited potential for disease control to better quantify the association between cholera and weather and understand their underlying mechanisms, for instance for the seasonal targeting of oral cholera vaccination and punctual and more long-term interventions to improve WASH access. By showing current gaps and limitations in the literature, this work may help define the next generation of studies on cholera and weather patterns to guide public health policy in a changing climate.

## Data Availability

All data and code produced in the present study will be made available on github upon publication.

