## Supplementary Material for "The association between cholera and weather: a systematic review and direction-of-effect meta-analysis"

#### Table of Contents

**1. Systematic review** **2**
1.1 Data Extraction 2
1.2 Location-specific data extraction 2
**2. Meta-regression analysis** **2**
**3. Supplementary Figures** **4**
**4. Supplementary Tables** **10**
**References** **22**

### 1. Systematic review

#### 1.1 Data Extraction

To enhance consistency and comparability, extracted variables were transformed as follows: all time-related variables, including temporal scales of exposure and outcome measurements, as well as lag times, were standardized to a uniform unit of days, assuming that one month is equal to 30 days and one season is equal to 90 days. They were then categorized into: 0 day, 1-14 days, 15-30 days, 31-365 days, >365 days and missing. For ratio effect estimates associated with decreasing exposures, both the estimates and their uncertainties were inverted. Association measurements reported in a logarithmic scale were converted back to a normal scale to facilitate interpretation and analysis.

#### 1.2 Location-specific data extraction

We obtained shapefiles for each study location from a global cholera incidence database and The Humanitarian Data Exchange (HDX) (1,2). For study areas comprising multiple locations, we merged their individual shapefiles into a single unified polygon. Locations were further categorized as subnational, national, and regional (e.g., spanning multiple countries). Locations within the 100 kilometers (KM) of the nearest ocean were classified as coastal; those beyond this threshold were classified as inland (3). Country-specific historical cholera status was characterized by the proportion of years with any World Health Organization (WHO)-reported cases between 2000-2023 (4,5). Country-years without reported cases were assumed to have zero incidence. Climatic zones were assigned using the Köppen-Geiger climate classification system, based on the zone covering the largest area within each study location (6). HDI values were obtained from the United Nation Development Progress (UNDP) Subnational HDI raster dataset (7). We computed the average HDI by aggregating values across the intersected areas between the HDI raster and the study location. HDI was then categorized using UNDP standards: 0-0.55 (low HDI), 0.55-0.7 (Medium HDI), 0.7-0.8 (High HDI), and 0.8-1 (Very high HDI) (8). We further classified locations as rural or urban based on population density, defining areas with fewer than 1,000 people per 1 $km^2$ as rural and those with 1,000 or more people per 1 $km^2$ as urban (9–11).

### 2. Meta-regression analysis

The aim of the meta-regression analysis was to assess the effect of relevant contextual factors on the direction of association between cholera and weather variables. To this end we used a Bayesian logistic regression model accounting both for estimate-level error as well as between study variability. We model the direction of effect *i* of study *s* as a binary variable $y_{i,s}$ (1 when association direction is positive, 0 if it is negative) which is a function of covariates encoded in matrix ***X*** as:

$$y_{i,s} \sim Binomial(\eta_{i,s}),$$
$$logit(\eta_{i,s}) = \alpha_s + X_{i,s}\beta + \epsilon_{i,s},$$

$$
\begin{aligned}
\alpha_s &\sim Normal(\mu_s, \sigma^S),\\
\epsilon_{i,s} &\sim Normal(0, \sigma),\\
\beta &\sim Normal(0, 0.5),\\
\sigma^S &\sim Normal(0, 1),\\
\sigma &\sim Normal(0, 1),
\end{aligned}
$$

where $\alpha_s$ is the study-level random effect, $\beta$ is the vector of covariate coefficients, and $\epsilon_{i,s}$ is the estimate-level error. We choose Normal priors for all parameters using recommended weakly informative priors for logistic regression.

### 3. Supplementary Figures

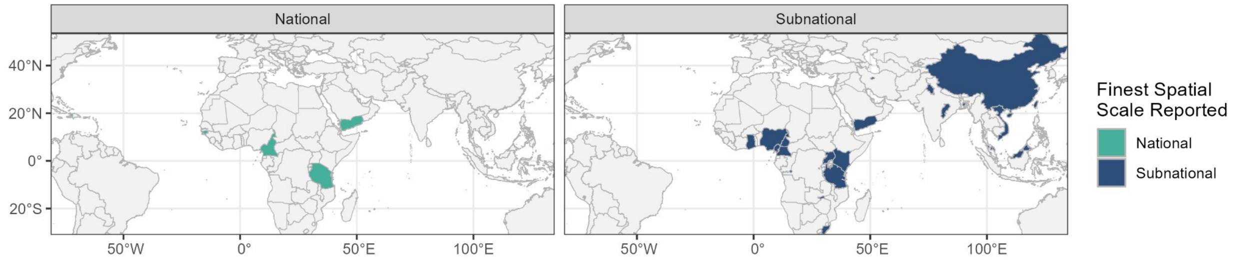


**Figure S1.** Geographic distribution of included studies at national and subnational scales, with colors representing the finest spatial scale reported in each study.

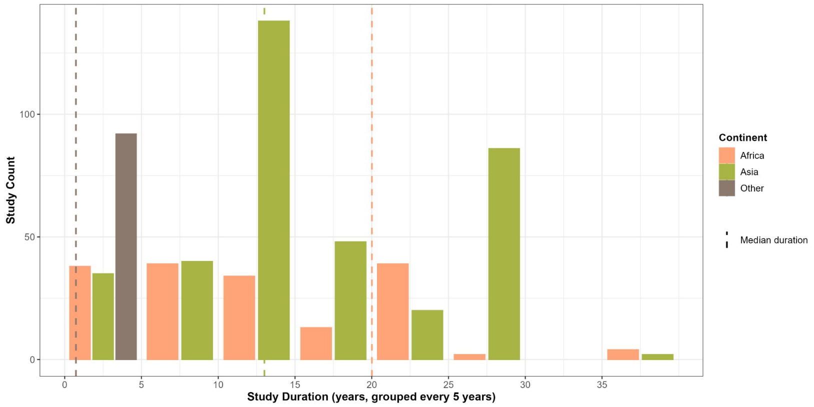


**Figure S2.** Distribution of study durations (years) by continent. Dashed lines represent the median study duration for each continent.

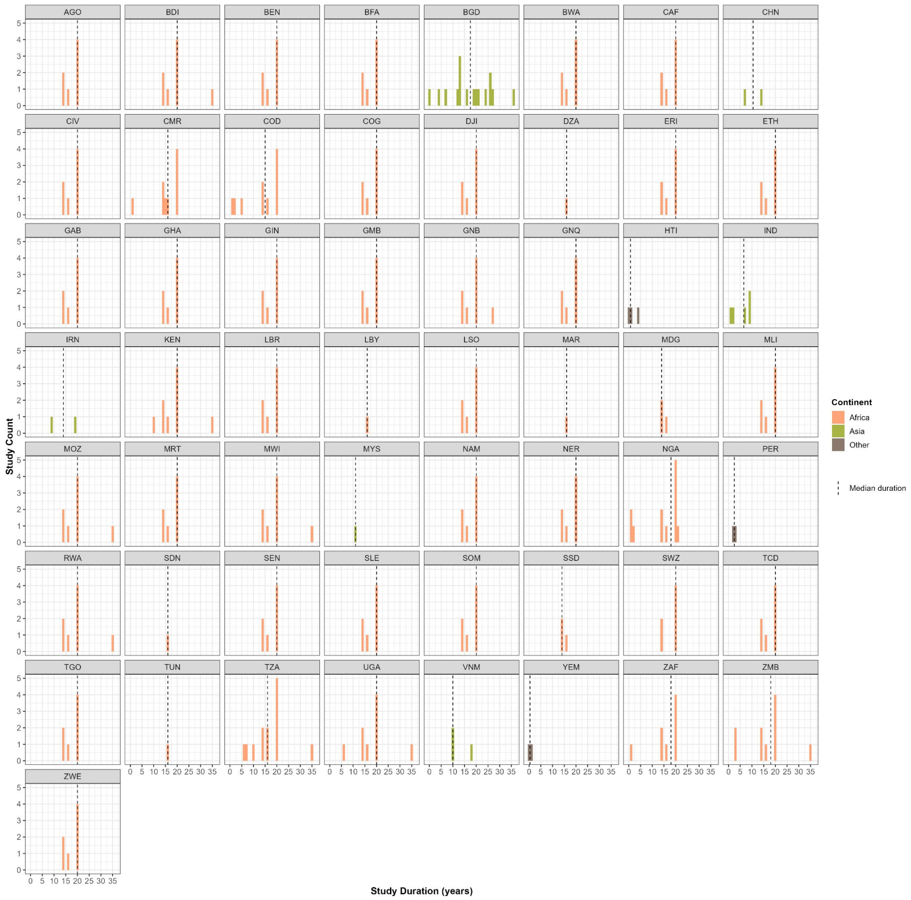


**Figure S3.** Distribution of study durations by country. Dashed lines represent the median study duration for each country. Colors represent different continents.

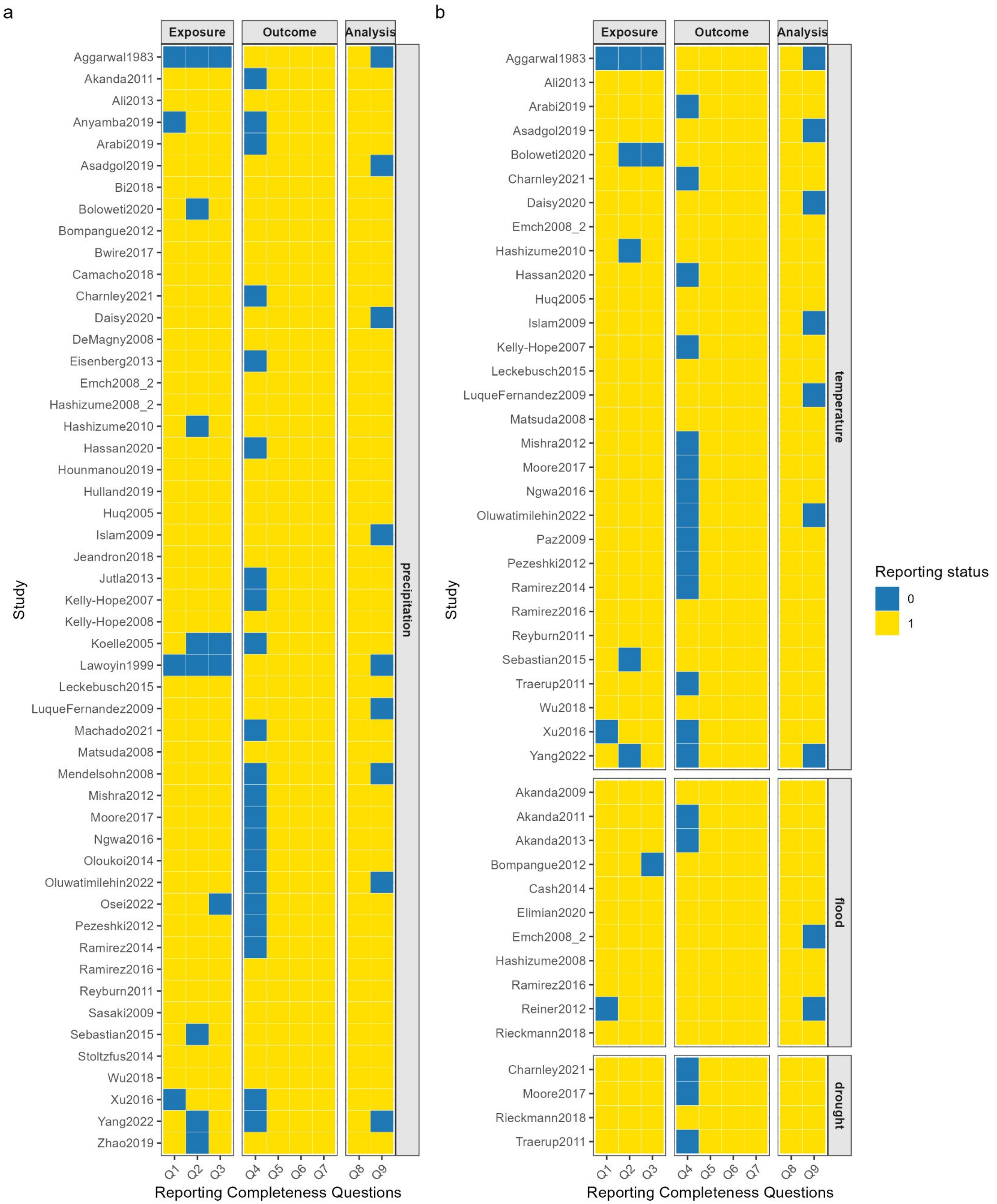


**Figure S4.** Study reporting completeness scoring tile plots by exposure group (i.e., precipitation, temperature, flood, and drought). Each row represents an individual study labeled with its unique study identifier. Columns correspond to questions used to assess the reporting quality, including exposure, outcome, and analysis-related aspects (see Table. S14). Variables are listed on the x axis. Colors represent the reporting status of each variable for each study.

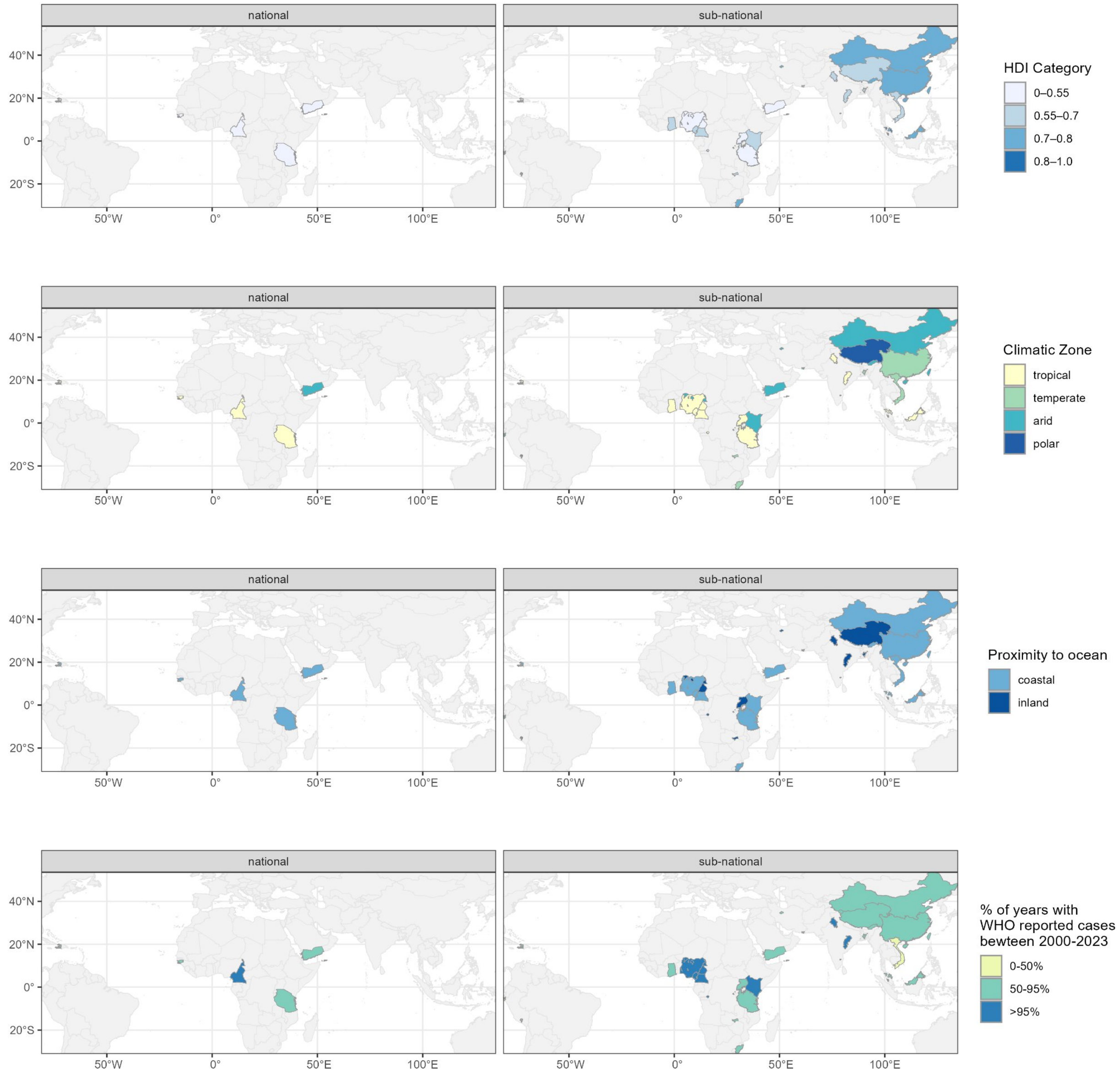


**Figure S5.** Geographic distributions of HDI, climatic zone, inland/coastal settings, and the percentage of years with WHO reported cases between 2000-2023 for national and subnational locations included in the studies.

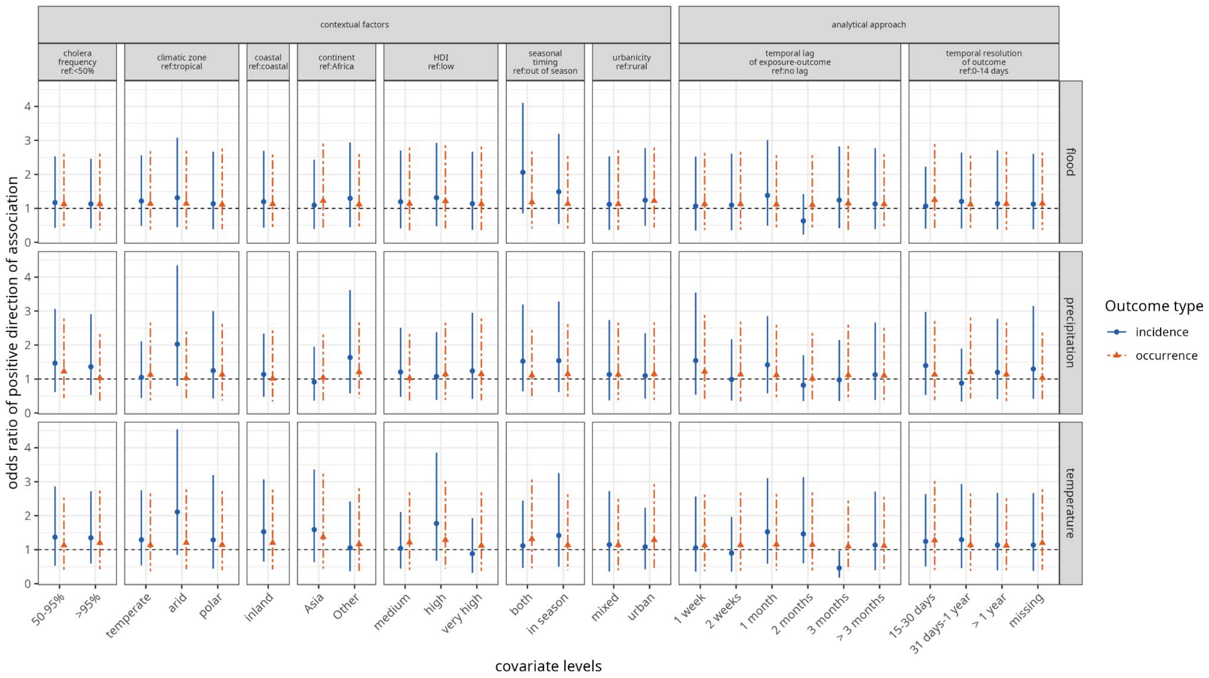


**Figure S6.** Meta-regression overall results.

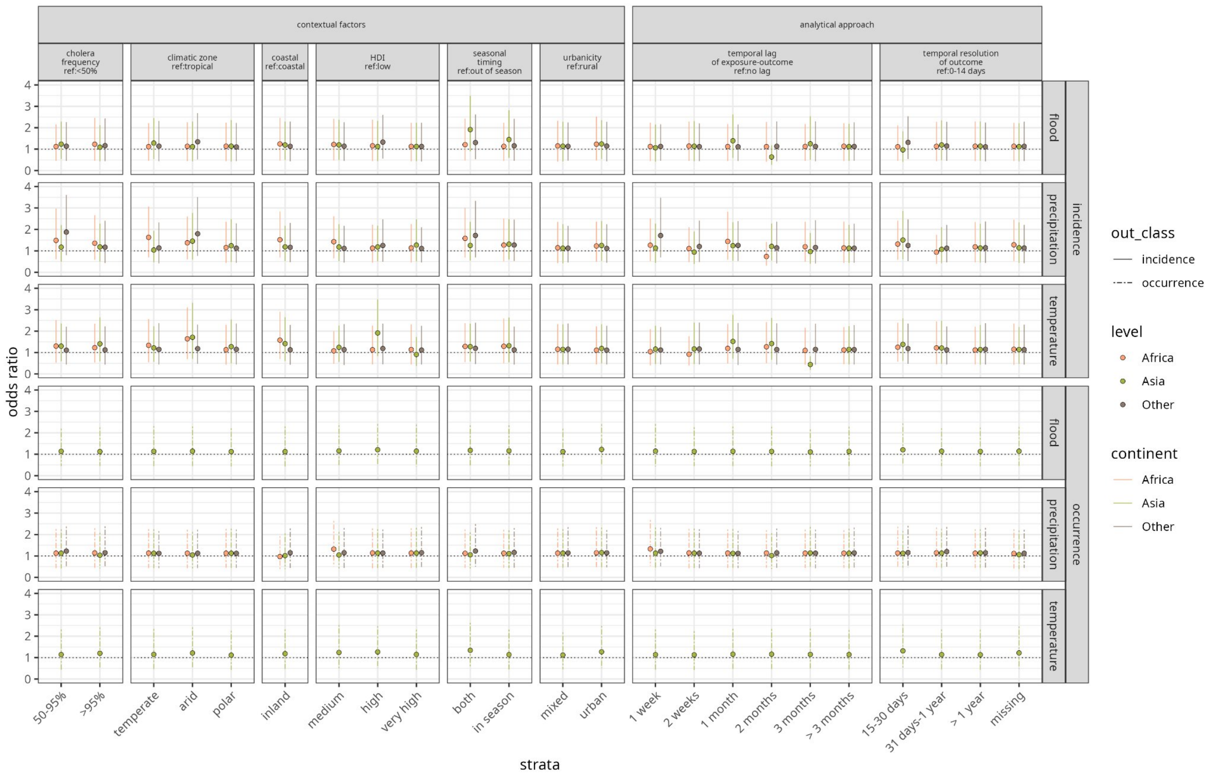


**Figure S7.** Meta-regression by continent

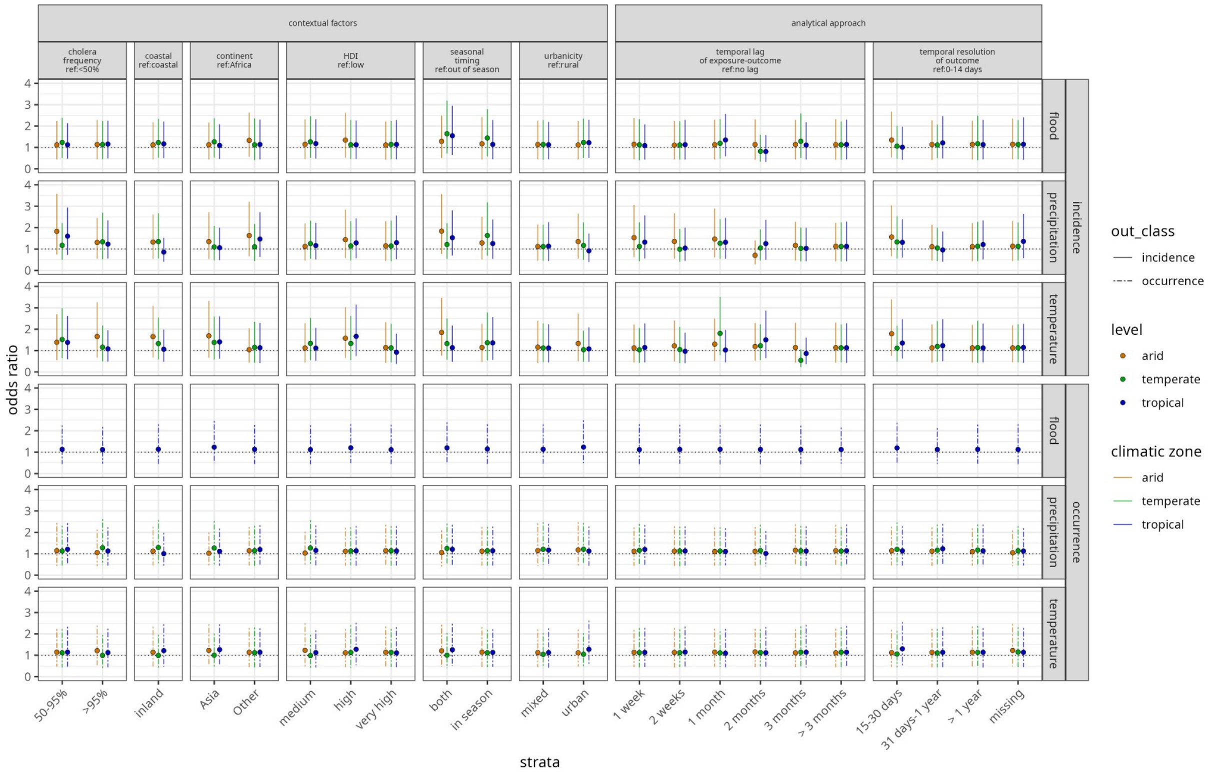


**Figure S8.** Meta-regression by climate zone.

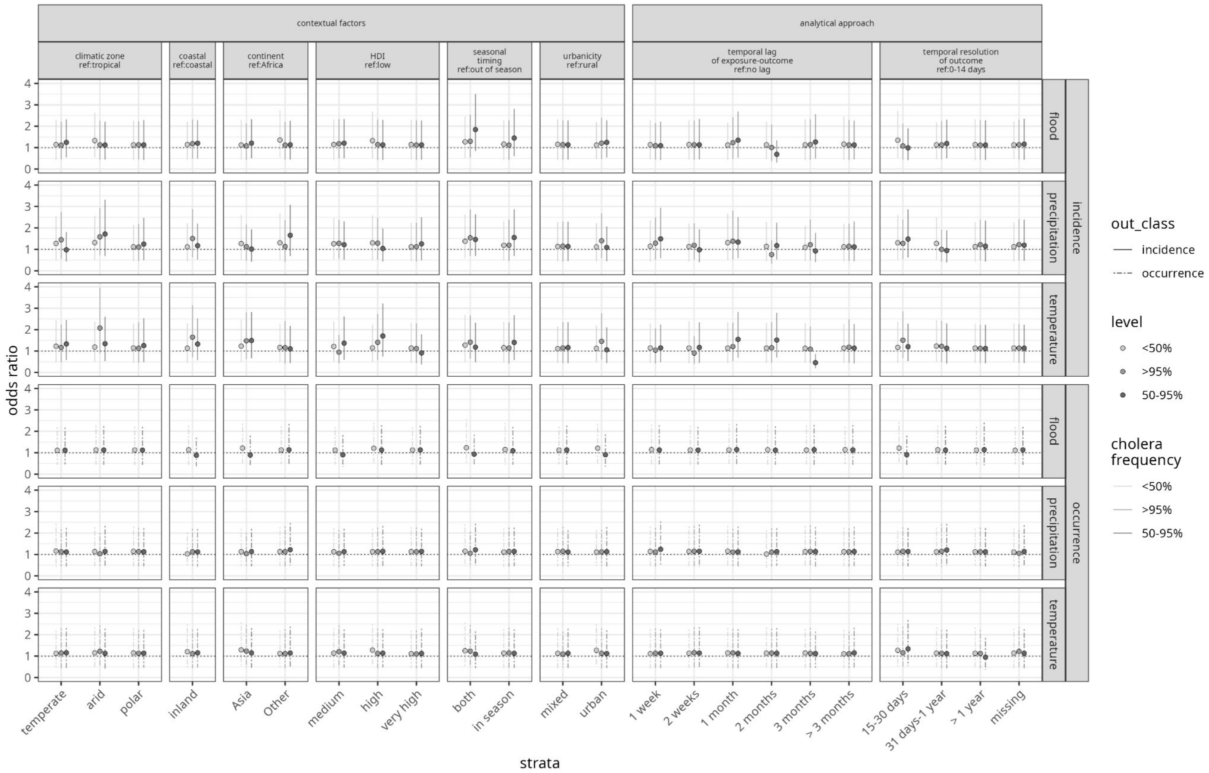


**Figure S9.** Meta-regression by frequency of cholera reporting to WHO.

### 4. Supplementary Tables

**Table S1.** Study characteristics across all studies

| Category | Characteristics | Precipitation | Temperature | Flood | Drought | Overall |
|---|---|---|---|---|---|---|
| **Total number of studies** | | 51 | 30 | 11 | 4 | 60 |
| **Spatial-temporal Characteristics** | | | | | | |
| **Study region** | **Africa** | | | | | |
| | **All countries** | 22 (43%) | 11 (37%) | 3 (27%) | 4 (100%) | 26 (41%) |
| | Democratic Republic of Congo | 5 (18%) | 3 (17%) | 2 (33%) | 3 (23%) | 6 (17%) |
| | Nigeria | 6 (21%) | 4 (22%) | 2 (33%) | 3 (23%) | 8 (22%) |
| | Tanzania | 6 (21%) | 5 (28%) | 1 (17%) | 4 (31%) | 9 (25%) |
| | Other countries | 11 (39%) | 6 (33%) | 1 (17%) | 3 (23%) | 13 (36%) |
| | **Asia** | | | | | |
| | **All countries** | 22 (43%) | 17 (57%) | 7 (64%) | 0 (0%) | 27 (45%) |
| | Bangladesh | 11 (48%) | 8 (44%) | 7 (78%) | 0 (0%) | 16 (55%) |
| | Other countries | 12 (52%) | 10 (56%) | 2 (22%) | 0 (0%) | 13 (45%) |
| | **Other continents** | | | | | |
| | **All countries** | 7 (14%) | 2 (7%) | 1 (9%) | 0 (0%) | 7 (12%) |
| | Haiti | 3 (43%) | 0 (0%) | 0 (0%) | 0 (0%) | 3 (43%) |
| | Peru | 2 (29%) | 2 (100%) | 1 (100%) | 0 (0%) | 2 (29%) |
| | Other countries | 2 (29%) | 0 (0%) | 0 (0%) | 0 (0%) | 2 (29%) |
| **Study spatial scale** | Subnational | 46 (87%) | 27 (87%) | 10 (91%) | 1 (25%) | 53 (85%) |
| | National | 5 (9%) | 1 (3%) | 0 (0%) | 0 (0%) | 5 (8%) |
| | Regional | 2 (4%) | 3 (10%) | 1 (9%) | 3 (75%) | 4 (7%) |
| **Study period** | <1 year | 10 (19%) | 3 (9%) | 3 (25%) | 0 (0%) | 13 (20%) |
| | 1-2 years | 3 (6%) | 1 (3%) | 0 (0%) | 0 (0%) | 3 (5%) |
| | 2-5 years | 9 (17%) | 6 (18%) | 1 (8%) | 0 (0%) | 9 (14%) |
| | 5-10 years | 11 (20%) | 7 (21%) | 1 (8%) | 1 (25%) | 14 (21%) |
| | >10 years | 21 (39%) | 17 (50%) | 7 (58%) | 3 (75%) | 27 (41%) |
| **% of years in 2000-2023 with WHO reported cholera cases** | 0 - 50% | 20 (35%) | 20 (36%) | 14 (31%) | 18 (36%) | 20 (35%) |
| | 50 - 95% | 28 (49%) | 26 (47%) | 22 (49%) | 22 (46%) | 28 (49%) |
| | > 95% | 9 (16%) | 9 (16%) | 9 (20%) | 8 (17%) | 9 (16%) |
| **Outcome Characteristics** | | | | | | |
| **Outcome type** | incidence | 42 (81%) | 26 (84%) | 8 (67%) | 2 (50%) | 49 (80%) |
| | occurrence | 4 (8%) | 3 (10%) | 2 (17%) | 2 (50%) | 5 (8%) |
| | other | 6 (12%) | 2 (7%) | 2 (17%) | 0 (0%) | 7 (11%) |
| **Temporal resolution** | 1-14 days | 13 (25%) | 5 (16%) | 2 (18%) | 0 (0%) | 14 (23%) |

| | | | | | | |
|---|---|---|---|---|---|---|
| **of outcome** | 15-30 days | 25 (49%) | 19 (61%) | 4 (36%) | 1 (25%) | 28 (46%) |
| | 31 days-1 year | 8 (16%) | 5 (16%) | 4 (36%) | 2 (50%) | 13 (21%) |
| | > 1 year | 2 (4%) | 1 (3%) | 1 (9%) | 1 (25%) | 3 (5%) |
| | missing | 3 (6%) | 1 (3%) | 0 (0%) | 0 (0%) | 3 (5%) |
| **Case definition** | Suspected | 3 (6%) | 1 (3%) | 0 (0%) | 0 (0%) | 3 (5%) |
| | Confirmed | 13 (24%) | 11 (34%) | 3 (25%) | 0 (0%) | 15 (23%) |
| | Deaths | 0 (0%) | 0 (0%) | 1 (8%) | 0 (0%) | 1 (2%) |
| | Mixed | 18 (33%) | 7 (22%) | 5 (42%) | 2 (50%) | 21 (33%) |
| | Unknown | 20 (37%) | 13 (41%) | 3 (25%) | 2 (50%) | 24 (38%) |
| **Statistical Methods** | | | | | | |
| **Measure of association effect** | Ratio | 23 (39%) | 16 (42%) | 4 (33%) | 4 (100%) | 30 (42%) |
| | Correlation coefficient | 28 (47%) | 16 (42%) | 6 (50%) | 0 (0%) | 32 (45%) |
| | Risk difference | 5 (9%) | 3 (8%) | 1 (8%) | 0 (0%) | 6 (9%) |
| | Attributable risk | 2 (3%) | 2 (5%) | 0 (0%) | 0 (0%) | 2 (3%) |
| | Other | 1 (2%) | 1 (3%) | 1 (8%) | 0 (0%) | 1 (1%) |
| **Reporting Completeness Score** | | | | | | |
| **Overall score*** | 9 (maximum) | 20 (39%) | 8 (27%) | 6 (55%) | 1 (25%) | 23 (38%) |
| | 5-8 | 31 (61%) | 22 (73%) | 5 (45%) | 3 (75%) | 37 (62%) |
| **Exposure-related score*** | 3 (maximum) | 40 (78%) | 24 (80%) | 9 (82%) | 4 (100%) | 47 (78%) |
| | <3 | 11 (22%) | 6 (20%) | 2 (18%) | 0 (0%) | 13 (22%) |
| **Outcome-related score*** | 4 (maximum) | 30 (59%) | 16 (53%) | 9 (82%) | 1 (25%) | 36 (60%) |
| | <4 | 21 (41%) | 16 (47%) | 2 (18%) | 3 (75%) | 24 (40%) |
| **Association-related score*** | 2 (maximum) | 42 (82%) | 23 (77%) | 9 (82%) | 4 (100%) | 49 (82%) |
| | <2 | 9 (18%) | 7 (23%) | 2 (18%) | 0 (0%) | 11 (18%) |

**Table S2.** Raw values for the composite category in each variable

| Variable | Category | Raw values | N studies |
|---|---|---|---|
| Measure of association effect | other | Probit model coefficient with non-linear association relationship | 1 |
| Outcome | other | Variation of monthly cholera cases | 1 |
| | | Cholera-related deaths | 2 |
| | | Whether an individual had a confirmed cholera infection resulting in hospital admission versus matched control period without cholera | 1 |
| | | Probability of being tested positive | 1 |
| | | High vs. Low risk of disease period | 1 |
| | | Ranking of monthly cholera cases among the same calendar month of other years | 1 |
| | | Whether the lower bound of 95% credible interval of daily reproductive number at the shehia level greater than 1 | 1 |
| Case definition | Mixed | Sero-incidence infections and bacteriologically confirmed cases | 1 |
| | | Sero-incidence infections, bacteriologically confirmed cases and suspected cases | 1 |
| | | Suspected and bacteriologically confirmed cases only | 1 |

**Table S3.** Study characteristics in Africa

| Category | Characteristics | Precipitation | Temperature | Flood | Drought | Overall |
|---|---|---|---|---|---|---|
| **Total number of studies** | | 22 | 11 | 3 | 4 | 26 |
| **Spatial-temporal Characteristics** | | | | | | |
| **Study region** | Central Africa | 7 (24%) | 5 (26%) | 2 (33%) | 3 (21%) | 8 (22%) |
| | Eastern Africa | 8 (28%) | 5 (26%) | 1 (17%) | 4 (29%) | 11 (30%) |
| | Northern Africa | 1 (3%) | 1 (5%) | 0 (0%) | 1 (7%) | 1 (2.7%) |
| | Southern Africa | 5 (17%) | 4 (21%) | 1 (17%) | 3 (21%) | 7 (19%) |
| | Western Africa | 8 (28%) | 4 (21%) | 2 (33%) | 3 (21%) | 10 (27%) |
| **Study spatial scale** | Subnational | 18 (78%) | 8 (67%) | 2 (67%) | 1 (25%) | 20 (74%) |
| | National | 3 (13%) | 1 (8.3%) | 0 (0%) | 0 (0%) | 3 (11%) |
| | Regional | 2 (8.7%) | 3 (25%) | 1 (33%) | 3 (75%) | 4 (15%) |
| **Study period** | 1-2 years | 2 (9%) | 0 (0%) | 0 (0%) | 0 (0%) | 2 (7%) |
| | 2-5 years | 4 (18%) | 2 (18%) | 0 (0%) | 0 (0%) | 4 (15%) |
| | 5-10 years | 4 (18%) | 2 (18%) | 0 (0%) | 1 (25%) | 5 (19%) |
| | <1 year | 4 (18%) | 1 (9%) | 2 (67%) | 0 (0%) | 6 (22%) |
| | >10 years | 8 (36%) | 6 (55%) | 1 (33%) | 3 (75%) | 10 (37%) |
| **% of years with WHO reported cholera cases** | 0 - 50% | 18 (38%) | 18 (38%) | 12 (29%) | 18 (38%) | 18 (38%) |
| | 50 - 95% | 22 (46%) | 22 (46%) | 21 (51%) | 22 (46%) | 22 (46%) |
| | > 95% | 8 (17%) | 8 (17%) | 8 (20%) | 8 (17%) | 8 (17%) |
| **Outcome Characteristics** | | | | | | |
| **Outcome type** | incidence | 19 (86%) | 10 (91%) | 1 (33%) | 2 (50%) | 21 (81%) |
| | occurrence | 1 (14%) | 1 (9%) | 1 (33%) | 2 (50%) | 2 (8%) |
| | other | 2 (9%) | 0 (0%) | 1 (33%) | 0 (0%) | 3 (12%) |
| **Temporal resolution of outcome** | 1-14 days | 6 (27%) | 3 (25%) | 1 (33%) | 0 (0%) | 6 (22%) |
| | 15-30 days | 8 (36%) | 5 (42%) | 0 (0%) | 1 (25%) | 9 (33%) |
| | 31-365 days | 5 (23%) | 4 (33%) | 1 (33%) | 2 (50%) | 8 (30%) |
| | >365 days | 1 (4.5%) | 0 (0%) | 1 (33%) | 1 (25%) | 2 (7%) |
| | missing | 2 (9%) | 0 (0%) | 0 (0%) | 0 (0%) | 2 (7%) |
| **Case definition** | Suspected | 2 (8.7%) | 1 (9%) | 0 (0%) | 0 (0%) | 2 (7%) |
| | Confirmed | 0 (0%) | 0 (0%) | 0 (0%) | 0 (0%) | 0 (0%) |
| | Deaths | 0 (0%) | 0 (0%) | 1 (33%) | 0 (0%) | 1 (4%) |
| | Mixed | 10 (43%) | 2 (18%) | 2 (67%) | 2 (50%) | 11 (41%) |
| | Unknown | 11 (48%) | 8 (73%) | 0 (0%) | 2 (50%) | 13 (48%) |
| **Statistical Methods** | | | | | | |
| **Measure of association effect** | Ratio | 12 (48%) | 8 (57%) | 3 (100%) | 4 (100%) | 17 (57%) |
| | Correlation coefficient | 11 (44%) | 4 (29%) | 0 (0%) | 0 (0%) | 11 (37%) |
| | Risk difference | 1 (4%) | 1 (7%) | 0 (0%) | 0 (0%) | 1 (3%) |
| | Attributable risk | 1 (4.0%) | 1 (7%) | 0 (0%) | 0 (0%) | 1 (3%) |
| | Other | 0 (0%) | 0 (0%) | 0 (0%) | 0 (0%) | 0 (0%) |
| **Any reported Uncertainty** | TRUE | 20 (91%) | 11 (100%) | 3 (100%) | 4 (100%) | 24 (92%) |
| | FALSE | 2 (9%) | 0 (0%) | 0 (0%) | 0 (0%) | 2 (8%) |
| **Reporting Completeness Score** | | | | | | |
| **Overall score** | 9 | 9 (41%) | 2 (18%) | 2 (67%) | 1 (25%) | 10 (38%) |
| | 5-8 | 13 (59%) | 9 (82%) | 1 (33%) | 3 (75%) | 16 (62%) |
| **Exposure-related score** | 3 | 18 (82%) | 10 (91%) | 2 (67%) | 4 (100%) | 21 (81%) |
| | <3 | 4 (18%) | 1 (9%) | 1 (33%) | 0 (0%) | 5 (19%) |
| **Outcome-related score** | 4 | 10 (45%) | 2 (18%) | 3 (100%) | 1 (25%) | 12 (46%) |
| | <4 | 12 (55%) | 9 (82%) | 0 (0%) | 3 (75%) | 14 (54%) |
| **Association-related score** | 2 | 18 (82%) | 9 (82%) | 3 (100%) | 4 (100%) | 22 (85%) |
| | <2 | 4 (18%) | 2 (18%) | 0 (0%) | 0 (0%) | 4 (15%) |

**Table S4.** Study characteristics in Asia

| Category | Characteristics | Precipitation | Temperature | Flood | Drought | Overall |
|---|---|---|---|---|---|---|
| **Total number of studies** | | 22 | 17 | 7 | 0 | 27 |
| **Spatial-temporal Characteristics** | | | | | | |
| **Study region** | Bangladesh | 7 (78%) | 11 (48%) | 8 (44%) | 0 (0%) | 16 (55%) |
| | China | 0 (0%) | 2 (9%) | 2 (11%) | 0 (0%) | 2 (7%) |
| | india | 1 (11%) | 4 (17%) | 3 (17%) | 0 (0%) | 5 (17%) |
| | Iran | 0 (0%) | 2 (9%) | 2 (11%) | 0 (0%) | 2 (7%) |
| | Malaysia | 0 (0%) | 1 (4%) | 1 (6%) | 0 (0%) | 1 (3%) |
| | Vietnam | 1 (11%) | 3 (13%) | 2 (11%) | 0 (0%) | 3 (10%) |
| **Study spatial scale** | Subnational | 22 (100%) | 17 (100%) | 7 (100%) | 0 (0%) | 27 (100%) |
| | National | 0 (0%) | 0 (0%) | 0 (0%) | 0 (0%) | 0 (0%) |
| | Regional | 0 (0%) | 0 (0%) | 0 (0%) | 0 (0%) | 0 (0%) |
| **Study period** | 1-2 years | 3 (5%) | 1 (3%) | 0 (0%) | 0 (0%) | 3 (4%) |
| | 2-5 years | 3 (12%) | 3 (14%) | 0 (0%) | 0 (0%) | 3 (9%) |
| | 5-10 years | 7 (28%) | 5 (24%) | 1 (13%) | 0 (0%) | 9 (28%) |
| | <1 year | 2 (8%) | 2 (10%) | 1 (13%) | 0 (0%) | 3 (9%) |
| | >10 years | 13 (52%) | 11 (52%) | 6 (75%) | 0 (0%) | 17 (53%) |
| **% of years with WHO reported cholera cases** | 0 - 50% | 1 (17%) | 1 (16.7%) | 1 (33%) | 0 (0%) | 1 (17%) |
| | 50 - 95% | 4 (67%) | 4 (66.7%) | 1 (33%) | 0 (0%) | 4 (67%) |
| | > 95% | 1 (17%) | 1 (16.7%) | 1 (33%) | 0 (0%) | 1 (17%) |
| **Outcome Characteristics** | | | | | | |
| **Outcome type** | incidence | 17 (74%) | 14 (78%) | 6 (75%) | 0 (0%) | 22 (79%) |
| | occurrence | 2 (9%) | 2 (11%) | 1 (13%) | 0 (0%) | 2 (7%) |
| | other | 4 (17%) | 2 (11%) | 1 (13%) | 0 (0%) | 4 (14%) |
| **Temporal resolution of outcome** | 1-14 days | 3 (14%) | 2 (12%) | 1 (14%) | 0 (0%) | 4 (15%) |
| | 15-30 days | 15 (68%) | 12 (71%) | 3 (43%) | 0 (0%) | 17 (63%) |
| | 31-365 days | 2 (9%) | 1 (6%) | 3 (43%) | 0 (0%) | 4 (15%) |
| | >365 days | 1 (5%) | 1 (6%) | 0 (0%) | 0 (0%) | 1 (4%) |
| | missing | 1 (5%) | 1 (6%) | 0 (0%) | 0 (0%) | 1 (4%) |
| **Case definition** | Suspected | 0 (0%) | 0 (0%) | 0 (0%) | 0 (0%) | 0 (0%) |
| | Confirmed | 13 (54%) | 11 (58%) | 3 (38%) | 0 (0%) | 15 (52%) |
| | Deaths | 0 (0%) | 0 (0%) | 0 (0%) | 0 (0%) | 0 (0%) |
| | Mixed | 3 (13%) | 2 (11%) | 2 (25%) | 0 (0%) | 5 (17%) |
| | Unknown | 8 (33%) | 6 (32%) | 3 (38%) | 0 (0%) | 9 (31%) |
| **Statistical Methods** | | | | | | |
| **Measure of association effect** | Ratio | 8 (31%) | 8 (36%) | 1 (13%) | 0 (0%) | 10 (30%) |
| | Correlation coefficient | 13 (50%) | 10 (45%) | 5 (63%) | 0 (0%) | 17 (52%) |
| | Risk difference | 3 (12%) | 2 (9%) | 1 (13%) | 0 (0%) | 4 (12%) |
| | Attributable risk | 1 (4%) | 1 (5%) | 0 (0%) | 0 (0%) | 1 (3%) |
| | Other | 1 (4%) | 1 (5%) | 1 (13%) | 0 (0%) | 1 (3%) |
| **Any reported uncertainty** | TRUE | 20 (91%) | 15 (88%) | 6 (86%) | 0 (0%) | 24 (89%) |
| | FALSE | 2 (9%) | 2 (12%) | 1 (14%) | 0 (0%) | 3 (11%) |
| **Reporting Completeness Score** | | | | | | |
| **Overall score** | 9 | 8 (36%) | 5 (29%) | 3 (43%) | 0 (0%) | 9 (33%) |
| | 5-8 | 14 (64%) | 12 (71%) | 4 (57%) | 0 (0%) | 18 (67%) |
| **Exposure-related score** | 3 | 16 (73%) | 12 (71%) | 6 (86%) | 0 (0%) | 20 (74%) |
| | <3 | 6 (27%) | 5 (29%) | 1 (14%) | 0 (0%) | 7 (26%) |
| **Outcome-related score** | 4 | 13 (59%) | 10 (59%) | 4 (57%) | 0 (0%) | 16 (59%) |
| | <4 | 9 (41%) | 7 (41%) | 3 (43%) | 0 (0%) | 11 (41%) |
| **Association-related score** | 2 | 17 (77%) | 12 (71%) | 5 (71%) | 0 (0%) | 15 (56%) |
| | <2 | 5 (23%) | 5 (29%) | 2 (29%) | 0 (0%) | 12 (44%) |

**Table S5.** Study characteristics in Other continents

| Category | Characteristics | Precipitation | Temperature | Flood | Drought | Overall |
|---|---|---|---|---|---|---|
| **Total number of studies** | | 7 | 2 | 1 | 0 | 7 |
| **Spatial-temporal Characteristics** | | | | | | |
| **Study region** | Haiti | 0 (0%) | 3 (43%) | 0 (0%) | 0 (0%) | 3 (43%) |
| | Peru | 1 (100%) | 2 (29%) | 2 (100%) | 0 (0%) | 2 (29%) |
| | Yemen | 0 (0%) | 2 (29%) | 0 (0%) | 0 (0%) | 2 (29%) |
| **Study spatial scale** | Subnational | 6 (75%) | 2 (100%) | 1 (100%) | 0 (0%) | 6 (75%) |
| | National | 2 (25%) | 0 (0%) | 0 (0%) | 0 (0%) | 2 (25%) |
| | Regional | 0 (0%) | 0 (0%) | 0 (0%) | 0 (0%) | 0 (0%) |
| **Study period** | 1-2 years | 1 (14%) | 1 (50%) | 0 (0%) | 0 (0%) | 1 (14%) |
| | 2-5 years | 2 (29%) | 1 (50%) | 1 (100%) | 0 (0%) | 2 (29%) |
| | 5-10 years | 0 (0%) | 0 (0%) | 0 (0%) | 0 (0%) | 0 (0%) |
| | <1 year | 4 (57%) | 0 (0%) | 0 (0%) | 0 (0%) | 4 (57% |
| | >10 years | 0 (0%) | 0 (0%) | 0 (0%) | 0 (0%) | 0 (0%) |
| **% of years with WHO reported cholera cases** | 0 - 50% | 1 (33%) | 1 (100%) | 1 (100%) | 0 (0%) | 1 (33%) |
| | 50 - 95% | 2 (67%) | 0 (0%) | 0 (0%) | 0 (0%) | 2 (67%) |
| | > 95% | 0 (0%) | 0 (0%) | 0 (0%) | 0 (0%) | 0 (0%) |
| **Outcome Characteristics** | | | | | | |
| **Outcome type** | incidence | 6 (86%) | 2 (100%) | 1 (100%) | 0 (0%) | 6 (86%) |
| | occurrence | 1 (14%) | 0 (0%) | 0 (0%) | 0 (0%) | 1 (14%) |
| | other | 0 (0%) | 0 (0%) | 0 (0%) | 0 (0%) | 0 (0%) |
| **Temporal resolution of outcome** | 1-14 days | 4 (57%) | 0 (0%) | 0 (0%) | 0 (0%) | 4 (57%) |
| | 15-30 days | 2 (29%) | 2 (100%) | 1 (100%) | 0 (0%) | 2 (29%) |
| | 31-365 days | 1 (14%) | 0 (0%) | 0 (0%) | 0 (0%) | 1 (14%) |
| | >365 days | 0 (0%) | 0 (0%) | 0 (0%) | 0 (0%) | 0 (0%) |
| | missing | 0 (0%) | 0 (0%) | 0 (0%) | 0 (0%) | 0 (0%) |
| **Case definition** | Suspected | 1 (14%) | 0 (0%) | 0 (0%) | 0 (0%) | 1 (14%) |
| | Confirmed | 0 (0%) | 0 (0%) | 0 (0%) | 0 (0%) | 0 (0%) |
| | Deaths | 0 (0%) | 0 (0%) | 0 (0%) | 0 (0%) | 0 (0%) |
| | Mixed | 4 (57%) | 2 (100%) | 1 (100%) | 0 (0%) | 4 (57%) |
| | Unknown | 2 (29%) | 0 (0%) | 0 (0%) | 0 (0%) | 2 (29%) |
| **Statistical Methods** | | | | | | |
| **Measure of association effect** | Ratio | 3 (38%) | 0 (0%) | 0 (0%) | 0 (0%) | 3 (38% |
| | Correlation coefficient | 4 (50%) | 2 (100%) | 1 (100%) | 0 (0%) | 4 (50%) |
| | Risk difference | 1 (13%) | 0 (0%) | 0 (0%) | 0 (0%) | 1 (13%) |
| | Attributable risk | 0 (0%) | 0 (0%) | 0 (0%) | 0 (0%) | 0 (0%) |
| | Other | 0 (0%) | 0 (0%) | 0 (0%) | 0 (0%) | 0 (0%) |
| **Any reported uncertainty** | TRUE | 7 (100%) | 2 (100%) | 1 (100%) | 0 (0%) | 7 (100%) |
| | FALSE | 0 (0%) | 0 (0%) | 0 (0%) | 0 (0%) | 0 (0%) |
| **Reporting Completeness Score** | | | | | | |
| **Overall score** | 9 | 3 (43%) | 1 (50%) | 1 (100%) | 0 (0%) | 3 (43%) |
| | 5-8 | 4 (57%) | 1 (50%) | 0 (0%) | 0 (0%) | 4 (57%) |
| **Exposure-related score** | 3 | 6 (86%) | 2 (100%) | 1 (100%) | 0 (0%) | 6 (86%) |
| | <3 | 1 (14%) | 0 (0%) | 0 (0%) | 0 (0%) | 1 (14%) |
| **Outcome-related score** | 4 | 3 (43%) | 1 (50%) | 0 (0%) | 0 (0%) | 4 (57%) |
| | <4 | 4 (57%) | 1 (50%) | 1 (100%) | 0 (0%) | 3 (43%) |
| **Association-related score** | 2 | 7 (100%) | 2 (100%) | 1 (100%) | 0 (0%) | 7 (100%) |
| | <2 | 0 (0%) | 0 (0%) | 0 (0%) | 0 (0%) | 0 (0%) |

**Table S6.** Incidence and precipitation by the direction of effect

| Category | Characteristics | Positive | Non-positive | P-value |
|---|---|---|---|---|
| **Total number of studies** | | 32 | 10 | - |
| **Spatial-temporal Characteristics** | | | | |
| **Study region** | Asia | 10 (31%) | 7 (70%) | 0.068 |
| | Africa | 16 (50%) | 3 (30%) | |
| | Other | 6 (19%) | 0 (0%) | |
| **Study spatial scale** | Subnational | 29 (85%) | 9 (90%) | 0.087 |
| | National | 5 (15%) | 0 (0%) | |
| | Regional | 0 (0%) | 1 (10%) | |
| **Study period** | 1-2 years | 1 (3%) | 1 (10%) | 0.15 |
| | 2-5 years | 8 (24%) | 1 (10%) | |
| | 5-10 years | 5 (15%) | 4 (40%) | |
| | <1 year | 9 (24%) | 0 (0%) | |
| | >10 years | 12 (35%) | 4 (40%) | |
| **% of years with WHO reported cholera cases** | 0 - 50% | 2 (11%) | 14 (30%) | 0.30 |
| | 50 - 95% | 11 (61%) | 24 (51%) | |
| | > 95% | 5 (28%) | 8 (19%) | |
| **Outcome Characteristics** | | | | |
| **Temporal resolution of outcome** | 0-14 days | 9 (28%) | 2 (20%) | 0.50 |
| | 15-30 days | 17 (53%) | 5 (50%) | |
| | 31 days-1 year | 3 (9%) | 3 (30%) | |
| | > 1 year | 1 (3%) | 0 (0%) | |
| | missing | 2 (6%) | 0 (0%) | |
| **Case definition** | Suspected | 3 (9%) | 0 (0%) | 0.12 |
| | Confirmed | 6 (18%) | 5 (50%) | |
| | Deaths | 0 (0%) | 0 (0%) | |
| | Mixed | 12 (35%) | 1 (10%) | |
| | Unknown | 13 (38%) | 4 (40%) | |
| **Statistical Methods** | | | | |
| **Measure of association effect** | Ratio | 12 (30%) | 5 (50%) | 0.60 |
| | Correlation coefficient | 22 (55%) | 4 (40%) | |
| | Risk difference | 4 (10%) | 1 (10%) | |
| | Attributable risk | 2 (5%) | 0 (0%) | |
| | Other | 0 (0%) | 0 (0%) | |
| **Any reported uncertainty** | TRUE | 29 (91%) | 9 (90%) | > 0.9 |
| | FALSE | 3 (9%) | 1 (10%) | |
| **Reporting Completeness Score** | | | | |
| **Overall score** | 9 | 10 (31%) | 5 (50%) | 0.50 |
| | 5-8 | 22 (69%) | 5 (50%) | |
| | ≤ 4 | 0 (0%) | 0 (0%) | |
| **Exposure-related score** | 3 | 23 (72%) | 9 (90%) | 0.50 |
| | <3 | 9 (28%) | 1 (10%) | |
| **Outcome-related score** | 4 | 19 (59%) | 6 (60%) | > 0.9 |
| | <4 | 12 (41%) | 4 (40%) | |
| **Association-related score** | 2 | 24 (75%) | 9 (90%) | 0.60 |
| | <2 | 8 (25%) | 1 (10%) | |

**Table S7.** Incidence and temperature by the direction of effect

| Category | Characteristics | Positive | Non-positive | P-value |
|---|---|---|---|---|
| **Total number of studies** | | 20 | 6 | - |
| **Spatial-temporal Characteristics** | | | | |
| **Study region** | Asia | 11 (55%) | 3 (50%) | 0.60 |
| | Africa | 8 (40%) | 2 (33%) | |
| | Other | 1 (5.0%) | 1 (17%) | |
| **Study spatial scale** | Subnational | 18 (90%) | 6 (86%) | 0.20 |
| | National | 0 (0%) | 1 (14%) | |
| | Regional | 2 (10%) | 0 (0%) | |
| **Study period** | 1-2 years | 1 (5%) | 0 (0%) | 0.40 |
| | 2-5 years | 4 (19%) | 2 (25%) | |
| | 5-10 years | 2 (10%) | 3 (38%) | |
| | <1 year | 2 (10%) | 1 (13%) | |
| | >10 years | 12 (57%) | 2 (25%) | |
| **% of years with WHO reported cholera cases** | 0 - 50% | 16 (32%) | 1 (17%) | 0.60 |
| | 50 - 95% | 25 (50%) | 3 (50%) | |
| | > 95% | 9 (18%) | 2 (33%) | |
| **Outcome Characteristics** | | | | |
| **Temporal resolution of outcome** | 0-14 days | 4 (19%) | 1 (17%) | 0.50 |
| | 15-30 days | 13 (62%) | 5 (83%) | |
| | 31 days-1 year | 4 (19%) | 0 (0%) | |
| | > 1 year | 0 (0%) | 0 (0%) | |
| | missing | 0 (0%) | 0 (0%) | |
| **Case definition** | Suspected | 1 (5%) | 0 (0%) | 0.70 |
| | Confirmed | 7 (33%) | 2 (33%) | |
| | Deaths | 0 (0%) | 0 (0%) | |
| | Mixed | 3 (14%) | 2 (33%) | |
| | Unknown | 10 (48%) | 2 (33%) | |
| **Statistical Methods** | | | | |
| **Measure of association effect** | Ratio | 10 (37%) | 3 (43%) | 0.70 |
| | Correlation coefficient | 12 (44%) | 4 (57%) | |
| | Risk difference | 3 (11%) | 0 (0%) | |
| | Attributable risk | 2 (7%) | 0 (0%) | |
| | Other | 0 (0%) | 0 (0%) | |
| **Any reported uncertainty** | TRUE | 18 (90%) | 6 (100%) | > 0.9 |
| | FALSE | 2 (10%) | 0 (0%) | |
| **Reporting Completeness Score** | | | | |
| **Overall score** | 9 | 3 (15%) | 3 (50%) | 0.20 |
| | 5-8 | 17 (85%) | 3 (50%) | |
| **Exposure-related score** | 3 | 16 (80%) | 4 (67%) | 0.90 |
| | <3 | 4 (20%) | 2 (33%) | |
| **Outcome-related score** | 4 | 10 (50%) | 4 (67%) | 0.80 |
| | <4 | 10 (50%) | 2 (33%) | |
| **Association-related score** | 2 | 7 (35%) | 6 (100%) | 0.20 |
| | <2 | 13 (65%) | 0 (0%) | |

**Table S8.** Incidence and flood by the direction of effect

| Category | Characteristics | Positive | Non-positive | P-value |
|---|---|---|---|---|
| **Total number of studies** | | 6 | 2 | - |
| **Spatial-temporal Characteristics** | | | | |
| **Study region** | Asia | 4 (57%) | 2 (100%) | 0.60 |
| | Africa | 1 (17%) | 0 (0%) | |
| | Other | 1 (17%) | 0 (0%) | |
| **Study spatial scale** | Subnational | 6 (100%) | 2 (100%) | > 0.9 |
| | National | 0 (0%) | 0 (0%) | |
| | Regional | 0 (0%) | 0 (0%) | |
| **Study period** | 1-2 years | 0 (0%) | 0 (0%) | 0.30 |
| | 2-5 years | 1 (17%) | 0 (0%) | |
| | 5-10 years | 0 (0%) | 1 (33%) | |
| | <1 year | 2 (33%) | 0 (0%) | |
| | >10 years | 3 (50%) | 2 (67%) | |
| **% of years with WHO reported cholera cases** | 0 - 50% | 1 (33%) | 0 (0%) | 0.70 |
| | 50 - 95% | 1 (33%) | 1 (50%) | |
| | > 95% | 1 (33%) | 1 (50%) | |
| **Outcome Characteristics** | | | | |
| **Temporal resolution of outcome** | 0-14 days | 2 (33%) | 0 (0%) | 0.60 |
| | 15-30 days | 2 (33%) | 1 (50%) | |
| | 31 days-1 year | 2 (33%) | 1 (50%) | |
| | > 1 year | 0 (0%) | 0 (0%) | |
| | missing | 0 (0%) | 0 (0%) | |
| **Case definition** | Suspected | 0 (0%) | 0 (0%) | 0.018 |
| | Confirmed | 2 (33%) | 0 (0%) | |
| | Deaths | 0 (0%) | 0 (0%) | |
| | Mixed | 4 (67%) | 0 (0%) | |
| | Unknown | 0 (0%) | 2 (100%) | |
| **Statistical Methods** | | | | |
| **Measure of association effect** | Ratio | 2 (33%) | 0 (0%) | > 0.9 |
| | Correlation coefficient | 4 (67%) | 2 (100%) | |
| | Risk difference | 0 (0%) | 0 (0%) | |
| | Attributable risk | 0 (0%) | 0 (0%) | |
| | Other | 0 (0%) | 0 (0%) | |
| **Any reported uncertainty** | TRUE | 5 (83%) | 2 (100%) | > 0.9 |
| | FALSE | 1 (17%) | 0 (0%) | |
| **Reporting Completeness Score** | | | | |
| **Overall score** | 9 | 4 (67%) | 0 (0%) | 0.40 |
| | 5-8 | 2 (33%) | 2 (100%) | |
| **Exposure-related score** | 3 | 4 (67%) | 2 (100%) | > 0.9 |
| | <3 | 2 (33%) | 0 (0%) | |
| **Outcome-related score** | 4 | 6 (100%) | 0 (0%) | 0.059 |
| | <4 | 0 (0%) | 2 (100%) | |
| **Association-related score** | 2 | 5 (83%) | 2 (100%) | > 0.9 |
| | <2 | 1 (17%) | 0 (0%) | |

**Table S9.** Vote counting summary by outcome type and effect direction for included studies

| Exposure | Outcome | Studies with overall positive effect direction | Studies with overall negative effect direction | Studies with inconclusive directionality | Overall direction | P-value |
|---|---|---|---|---|---|---|
| Precipitation | Incidence | 32 | 7 | 3 | positive | < 0.001 |
| | Occurrence | 0 | 2 | 2 | negative | 0.29 |
| | Other | 3 | 2 | 1 | positive | 0.82 |
| Temperature | Incidence | 20 | 0 | 6 | positive | < 0.001 |
| | Occurrence | 2 | 0 | 1 | positive | 0.30 |
| | Other | 1 | 0 | 1 | positive | 0.63 |
| Flood | Incidence | 6 | 0 | 2 | positive | 0.021 |
| | Occurrence | 2 | 0 | 0 | positive | 0.50 |
| | Other | 1 | 1 | 0 | positive | >0.9 |
| Drought | Incidence | 0 | 1 | 1 | negative | 0.63 |
| | Occurrence | 1 | 1 | 0 | positive | > 0.9 |

**Table S10.**Vote counting summary by outcome type and effect direction among studies with complete reporting status

| Exposure | Outcome | Studies with overall positive effect direction | Studies with overall negative effect direction | Studies with inconclusive directionality | Overall direction | P-value |
|---|---|---|---|---|---|---|
| Precipitation | Incidence | 10 | 3 | 2 | positive | 0.070 |
| | Occurrence | 0 | 0 | 1 | no direction | - |
| | Other | 3 | 1 | 1 | positive | 0.46 |
| Temperature | Incidence | 3 | 0 | 3 | positive | 0.28 |
| | Occurrence | 1 | 0 | 0 | positive | > 0.9 |
| | Other | 1 | 0 | 1 | positive | > 0.9 |
| Flood | Incidence | 4 | 0 | 0 | positive | 0.13 |
| | Occurrence | 2 | 0 | 0 | positive | 0.50 |
| | Other | 1 | 0 | 0 | positive | > 0.9 |
| Drought | Occurrence | 1 | 0 | 0 | positive | > 0.9 |

**Table S11.** Vote counting summary by outcome type and effect direction for studies in Africa

| Exposure | Outcome | Studies with overall positive effect direction | Studies with overall negative effect direction | Studies with inconclusive directionality | Overall direction | P-value |
|---|---|---|---|---|---|---|
| Precipitation | Incidence | 16 | 2 | 1 | positive | < 0.001 |
| | Occurrence | 0 | 1 | 0 | negative | > 0.9 |
| | Other | 1 | 1 | 0 | positive | > 0.9 |
| Temperature | Incidence | 8 | 0 | 2 | positive | 0.006 |
| | Occurrence | 0 | 0 | 1 | no direction | - |
| Flood | Incidence | 1 | 0 | 0 | positive | > 0.9 |
| | Occurrence | 1 | 0 | 0 | positive | > 0.9 |
| | Other | 1 | 0 | 0 | positive | > 0.9 |
| Drought | Incidence | 0 | 1 | 1 | negative | 0.63 |
| | Occurrence | 1 | 1 | 0 | positive | > 0.9 |

**Table S12.** Vote counting summary by outcome type and effect direction for studies in Asia

| Exposure | Outcome | Studies with overall positive effect direction | Studies with overall negative effect direction | Studies with inconclusive directionality | Overall direction | P-value |
|---|---|---|---|---|---|---|
| Precipitation | Incidence | 10 | 5 | 2 | positive | 0.25 |
| | Occurrence | 0 | 1 | 1 | negative | 0.63 |
| | Other | 2 | 1 | 1 | positive | 0.77 |
| Temperature | Incidence | 11 | 0 | 3 | positive | < 0.001 |
| | Occurrence | 2 | 0 | 0 | positive | 0.50 |
| | Other | 1 | 0 | 1 | positive | 0.63 |
| Flood | Incidence | 4 | 0 | 2 | positive | 0.077 |
| | Occurrence | 1 | 0 | 0 | positive | > 0.9 |
| | Other | 0 | 1 | 0 | negative | > 0.9 |

**Table S13.** Vote counting summary by outcome type and effect direction for studies in other continents

| Exposure | Outcome | Studies with overall positive effect direction | Studies with overall negative effect direction | Studies with inconclusive directionality | Overall direction | P-value |
|---|---|---|---|---|---|---|
| precipitation | Incidence | 6 | 0 | 0 | positive | 0.03 |
| | Occurrence | 0 | 0 | 1 | no direction | - |
| Temperature | Incidence | 1 | 0 | 1 | positive | 0.63 |
| Flood | Incidence | 1 | 0 | 0 | positive | > 0.9 |

**Table S14 Question summary of the Study Reporting Completeness Score System**

| Category | Question number and short title | Explanation |
|---|---|---|
| Exposure | Q1. Data source reported | Is the source of the weather exposure variable clearly specified in the study? |
| | Q2. Spatial scale reported | Is the spatial scale of the weather exposure variable (e.g., regional, national, provincial, etc) clearly specified in the study? |
| | Q3. Temporal scale reported | Is the temporal scale of the weather exposure variable (e.g., daily, weekly, monthly, etc) clearly specified in the study? |
| Outcome | Q4. Case definition reported | Does the study provide a clear definition for the cholera cases in the outcome variable (e.g., suspected cases, confirmed cases, cholera-related deaths, etc)? |
| | Q5. Case presentation reported | Does the study clearly describe how the cholera cases were presented, specifying whether the outcome variable includes only medically attended patients or also captures community-based cases? |
| | Q6. Spatial scale reported | Is the spatial scale of the cholera outcome variable (e.g., regional, national, provincial, etc) clearly specified in the study? |
| | Q7. Temporal scale reported | Is the temporal scale of the cholera outcome variable (e.g., daily, weekly, monthly, etc) clearly specified in the study? |
| Association | Q8. Effect measurements reported | Does the study clearly report a quantitative measurement of association (e.g., ratio, correlation coefficient, relative risk, etc)? |
| | Q9. Uncertainty reported | Does the study report statistical uncertainty for the association measurements? |

2. Website [Internet]. Available from: https://data.humdata.org/
